# Working in values-discrepant environments inhibits clinicians’ ability to provide compassion and reduces wellbeing: a cross-sectional study

**DOI:** 10.1101/2022.11.09.22282159

**Authors:** Alina Pavlova, Sarah-Jane Paine, Shane Sinclair, Anne O’Callaghan, Nathan S Consedine

## Abstract

**Objectives:** to describe the association between personal and organisational value discrepancies and compassion ability, burnout, job satisfaction, absenteeism, and consideration of early retirement among healthcare professionals.

**Design:** online cross-sectional survey.

**Setting:** primary, secondary, and tertiary care.

**Participants:** 1025 current practising clinicians (doctors, nurses, and allied health professionals) in Aotearoa/New Zealand.

**Main outcome measures:** The Sinclair Compassion Questionnaire – Healthcare Provider Ability and Competence Self-Assessment, The Maslach Burnout Inventory - Human Services Survey abbreviated 2-Question Summative Score, 10-item Warr-Cook-Wall Job Satisfaction questionnaire, measures of absenteeism and consideration of early retirement.

**Results:** Perceived discrepancies between personal and organisational values predicted lower compassion ability (*B* =-0.006, 95% CI [-0.01, -0.00], P<0.001, f2=0.05) but not competence (p=0.24), lower job satisfaction (*B* =-0.20, 95% CI [-0.23, -0.17], P<0.001, f2=0.14), higher burnout (*B* =0.02, 95% CI [0.01, 0.03], P<0.001, f2=0.06), absenteeism (*B* =0.004, 95% CI [0.00, 0.01], P=0.01 f2=0.01), and greater consideration of early retirement (*B* =0.02, 95% CI [0.00, 0.03], P=0.04, f2=0.004).

**Conclusions:** Working in value-discrepant environments predicts a range of poorer outcomes among healthcare professionals, including the ability to be compassionate. Scalable organisational and systems level interventions that address operational processes and practices that lead to the experience of value discrepancies are recommended to improve clinician performance and wellbeing outcomes.

**Study registration: the study was pre-registered on AsPredicted (Registration number 75407):** *What is already known on this topic:*

- Compassion predicts better patient outcomes and clinician quality of life
- Both personal and perceived organisational values predict variability in clinicians’ ability to show compassion and burnout
- Psychological tension associated with possibility of having to behave inconsistently with one’s own values, attitudes, and believes may result in unhelpful defence mechanisms associated with a range of negative outcomes

*What this paper adds:*

- Working in value-discrepant environments is associated with a lower ability to show compassion, lower job satisfaction, and higher burnout, absenteeism, and intention to retire early, even when overall competency is not affected
- Psychological tension and a low expectancy of positive outcomes seem likely to contribute to the link between being situated in value-discrepant environments and negative professional outcomes
- The findings of this study are non-consistent with the notion of compassion fatigue as reflecting the cost of caring that arises from exposure to repeated suffering. It seems more likely that not being able to practice compassionately due to conflicting personal-organisational values ultimately results in poorer professional wellbeing
- Organisational and fiscal level interventions that address operational processes and practices that lead to perceived value discrepancies are recommended and should be more effective for scalable improvement of health professional performance and wellbeing outcomes

## Background

Compassion is expected by patients, stipulated in medical codes of ethics, and morally mandated.^1–4^ Early data suggest that compassion predicts better patient outcomes ^5–7^ and professional’s quality of life.^5, 8–11^ However, patients report that their experiences of compassion not only vary, but are increasingly lacking in their healthcare experiences.^12–16^ The rates of professional burnout are also increasing,^17^ and absenteeism and early retirement – the costs associated with impaired professional quality of life – represent a serious burden to health systems worldwide.^18–20^

While notions of compassion fatigue have dominated discussions of compassion in health for some time ^21, 22^, it is increasingly recognised accepted that compassion is unlikely to cause fatigue ^22, 23^ and that an array of factors may interfere with the ability to express compassion.^8, 23–25^ However, while studies of compassion and compassion predictors are increasing,^5, 8, 9, 24, 26^ most research has focused on individual-level factors, notably factors that are fixed (e.g. speciality, gender) and may not be amenable to intervention.^24^ With the exception of a small number of qualitative studies,^27–29, 30^ little is known regarding how environmental or systems factors might impact clinicians’ ability to be compassionate.

Therefore, the primary aim of this large scale pre-registered study was to empirically test how organisational environments, in particular organisational cultures, may impact compassion and professional wellbeing.

The suggestion that organisational cultures impact care is not new.^31^ However, most accounts are theoretical and anecdotal rather than empirically based; robust methods by which to operationalise the potential effects of organisational culture have been missing.^32^ Prior studies suggest that both personal and organisational values – or (cultural) ideals to which both individuals and organisations strive ^33^ - predict compassion ^24^ and burnout ^34, 35^ and there is evidence that individuals who pursue healthcare careers are motivated by socially-focused values such as humility, compassion, and caring.^36–38^ Healthcare organisations are likely to signal similar humanistic values in public communications.^39–42^ However, little is known about how these values are actualized within these organisations, especially when fiscal austerity and uncertainty ^43^ coupled with the increased service demands exacerbated by the global Covid-19 pandemic ^44^ may have led to a “mission drift” where external pressures shift organisational vision, values, and goals from humanistic to more operational concerns.^45–47^ Again, some works imply that “business-oriented” environments that value efficiency, busy-ness, and emotional toughness can inhibit compassion.^28, 29, 48^ Of particular note, focusing on operational rather than socially-oriented values may conflict with personal values and beliefs.^36–38^ Theory suggests that engaging in behaviours that are inconsistent with personal beliefs can undermine personal and professional functioning (i.e. the theory of cognitive dissonance),^49^ create a ‘stress of conscience’,^50, 51^ and underpin moral distress.^52^ On this basis, we hypothesized that working in healthcare environments experienced as discrepant with one’s personal values would be associated with a lower ability to express compassion and lower job satisfaction and, conversely, with higher burnout, absenteeism, and intentions for early retirement.

## Methods

### Design

The study used a cross-sectional design and was conducted via an anonymous and voluntary online survey. This study was approved by the Auckland Health Research Ethics Committee on the 21st of October 2021 (Approval Number AH23221) and received locality approvals from each of 20 District Health Boards (DHBs) in Aotearoa/New Zealand (NZ). We adhered to The Strengthening the Reporting of Observational Studies in Epidemiology (STROBE) guidelines ^53^ and the Statistical Analyses and Methods in the Published Literature (SAMPL) guidelines ^54^ for cross-sectional studies in this report.

### Participants

English-speaking clinicians (medical doctors, nurses, and allied health professionals) aged over 18 years who were currently practicing, worked part-time or fulltime in NZ and had “regular contact with patients as a part of their job” were eligible to participate. A total of one thousand three hundred seventy-six people responded to the study advertisement with 1371 consenting. One hundred and twelve participants were excluded from the analysis due to not passing eligibility criteria (e.g. did not answer screening eligibility questions, not currently practicing, no clinical patient contact). Of 1259 eligible participants, 1025 provided sufficient data with regards to main predictor and represent this report’s sample (Figure 1).

**Figure 1:**
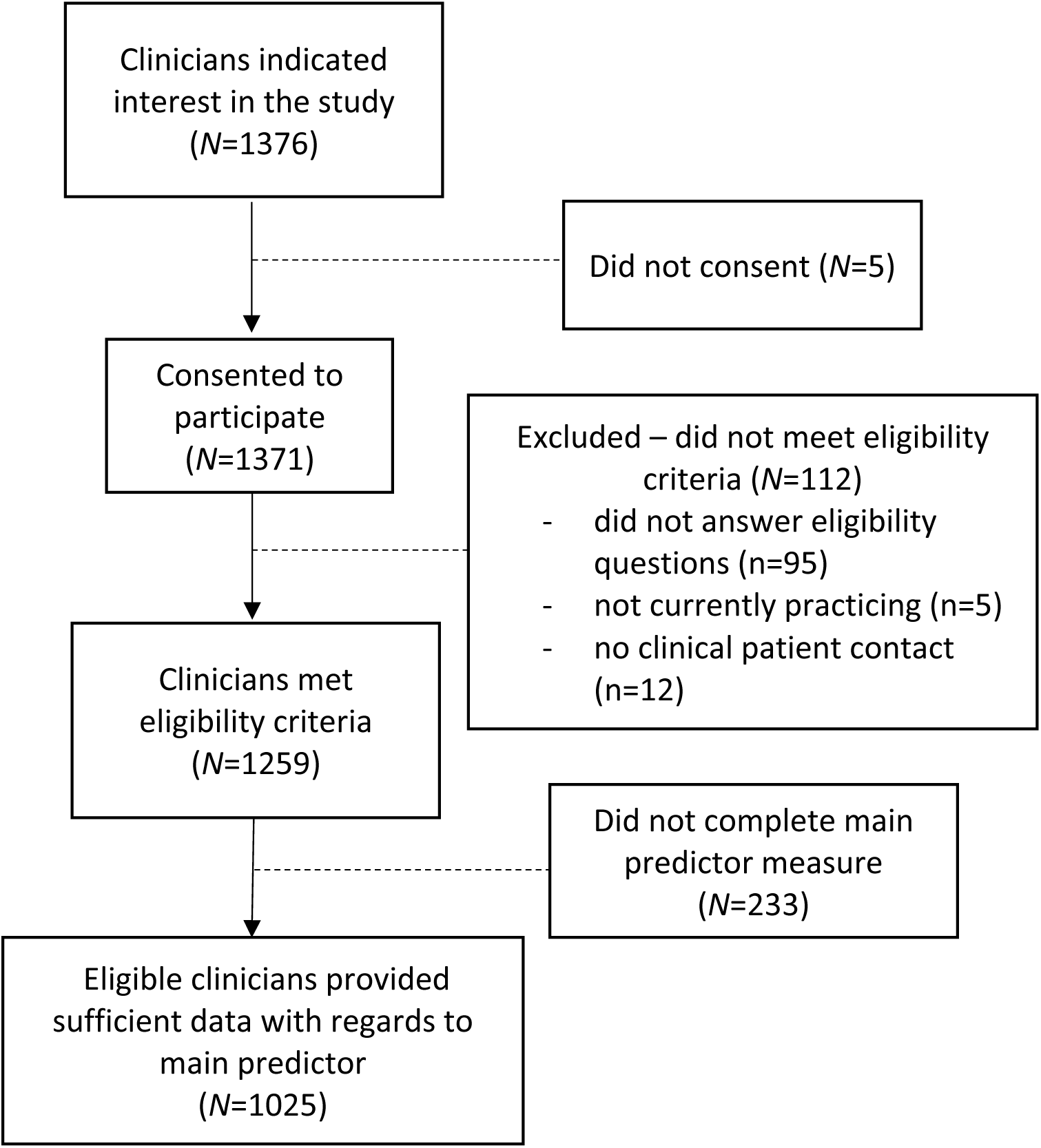
Flowchart of recruitment to final healthcare sample (N=1025)

### Bias and sample size

To avoid reporting bias, the study design, operationalisations, and hypotheses were pre-registered on AsPredicted (Registration number 75407: Hypothesis 1). Given core questions would be addressed using multiple regression analysis with approximately 6 predictors, pwr R^55^ suggested a sample size of 90 participants for a given category (e.g. occupation, ethnicity, etc) was needed to achieve 80% power with a confidence interval of 95% (estimated error of .05) and a medium size effect of f ²= 0.15. Thus, we aimed to recruit a minimum of 270 clinicians or 90 participants for each of the major professional groups in this context (doctors, nurses, allied health).

### Clinician involvement

Clinicians were involved in the design and conduct of this research, including coming up with the research question, choice of outcome measures, and methods of recruitment.

Once the study has been published, participants will be informed of the results via newsletter written in plain English.

### Procedures

Participants were recruited via DHBs employee newsletters and advertisements were widely shared via professional unions (doctors, nurses and allied health), Primary Health Organisations, and Kaupapa Māori and Hauora Māori Organisations (Indigenous healthcare organisations). Additionally, the call to participate was shared with health professional alumni from the University of Auckland and University of Otago – the two universities that train medical students. Recruitment took place from February to May 2022. As an incentive, participants could opt in to participate in a prize draw to win an iPad. Participation was voluntary and anonymous, with all participants having to first familiarise themselves with the study materials and sign an electronic consent form. The electronic survey took approximately 20 minutes to complete. The survey was titled ‘Institutional Barriers to care for kaimahi haoura (healthcare workers)’ to avoid self-selection for participation in compassion-related research.

### Measurements Primary outcomes

#### Compassion ability (SCQ-HCPASA)

Health professionals’ ability to express compassion was assessed by The Sinclair Compassion Questionnaire – Healthcare Provider Ability Self-Assessment adapted from the validated Sinclair Compassion Questionnaire (SCQ) that measures patients’ experiences of compassion.^56^ Clinicians were asked to rate how often they were able to show compassion in their work environment on a 5-point Likert scale ranging from 1= ‘Never able’ to 5= ‘Always able’. The scale indexing clinicians’ ability to show compassion consisted of 15-items from the original SCQ (e.g. “showing genuine concern”, “being attentive”, “really understanding patients’ needs”) focusing on patient perceptions of clinicians.

Given that the SCQ-HCPASA was only recently developed, a preliminary Exploratory Factor Analysis assessing the underlying structure of the SCQ-HCPASA measure was conducted in R (fa method). The traditional statistical assumptions were checked. The Kaiser–Meyer–Olkin (KMO) value was 0.96 and Bartlett’s Test of Sphericity was significant (χ2= 10,869.60 df=105, p<0.001), supporting the factorability of the correlation matrix. Consistent with screeplot inspection, parallel analysis ^57^ revealed a single component. Subsequent factor analysis with oblimin rotation and using Principle Axis Factoring method explained 57% of the variance, indicating a one factor model (χ2= 1,104.03, df=1025, p<0.001) (Appendix 1). Items were highly reliable (α = .95). The scale was positively correlated with Healthcare Provider Compassion Competency Self-Assessment (see below) (r=0.47, p<0.001) (see below) and with the Compassionate Love Scale (CLS-H-SF) (r=0.27, p<0.001)^58–60^ and general Self-Efficacy (r=0.34, p<0.001) scales.^61^

#### Compassion Competence (SCQ-HCPCSA)

Clinicians’ compassion competence was assessed by The Sinclair Compassion Questionnaire – Healthcare Provider Competence Self-Assessment, developed and measured similarly to SCQ-HCPASA. Clinicians were asked to rate how competent they felt in their compassion skills on the 5-point Likert scale ranging from 1= ‘Not at all competent’ to 5= ‘Completely competent’. The scale indexing the same 15-items from the original SCQ as SCQ-HCPASA.

Exploratory Factor Analysis was conducted in R (fa method), with the same parameters as SCQ-HCPASA. The Kaiser–Meyer–Olkin (KMO) value was 0.96 and Bartlett’s Test of Sphericity was significant (χ2= 9,065.23, df=105, p<0.001), supporting the factorability of the correlation matrix. Consistent with screeplot inspection, parallel analysis revealed a single component. Subsequent factor analysis explained 51% of the variance, indicating a one factor model (χ2= 904.43, df=1025, p<0.001) (Appendix 2). Items were highly reliable (α = .94). The scale was positively correlated the Compassionate Love Scale (CLS-H-SF) (r=0.31, p<0.001)^58–60^ and general Self-Efficacy scale (r=0.37, p<0.001).^61^

#### Burnout

To minimise participant attrition, burnout was assessed using The Maslach Burnout Inventory – Human Services Survey (MBI-HSS) abbreviated 2-Question Summative Score.^62^ Participants rated the following two items – “I feel burned out from my work” and “I have become more callous toward people since I took this job” – using a 1 (‘Never’) to 5 (‘Always’) Likert scale. In the original study, a summative score >3 demonstrated a sensitivity and specificity of 93.6% and 73.0% compared to the full MBI-HSS. This two-item burnout scale has good convergent validity, with correlations of 0.81 with Emotional Exhaustion and 0.73 with depersonalisation that are consistent with prior work^63, 64^ and better than that of the single-item measure.^65^

### Secondary outcomes

#### Job Satisfaction

Job satisfaction was assessed using a validated 10-item Warr-Cook- Wall questionnaire;^66, 67^ the scale is used extensively in healthcare populations.^68–72^ The first nine items of the scale index satisfaction with various aspects of the job (e.g. working conditions, autonomy, relationships with colleagues, etc), with the final item being an “overall” rating. Items are rated using a 7-item Likert scale with anchors ranging from 0=extremely dissatisfied to 7=extremely satisfied. In the original study, overall job satisfaction showed convergent validity with life satisfaction (*r*=0.42, *p*<0.05) and happiness (*r*=0.49, *p*<0.05) and was negatively correlated with anxiety (*r*=-0.24, *p*<0.05). A higher overall score indicates higher job satisfaction and total job satisfaction is the sum of the first nine items. In our sample the summative total score and overall satisfaction score were highly correlated (r=0.80, p<0.01), hence, the summative satisfaction score was used. Internal reliability for the first nine items was high (α = .88) in line with other studies (0.85-0.97).^66, 69, 70^

#### Absenteeism

Absenteeism was measured as a sum of self-reported number of days per year a person indicated being absent from work for either health-related, family-related, or burnout-related reasons, not including annual leave. A recent meta-analysis showed self-reported absenteeism was reliable over time (test-retest reliability of 0.79) and highly convergent with records (*r* = 0.73).^73^ It is, however, worth noting that the same meta-analysis also showed that employees tend to underreport their absences, with a mean difference of 2.03 days (SD = 2.19).^73^

#### Consideration of early retirement

Consideration of possible early retirement was measured by subtracting participant age from the legal NZ retirement age (currently 65 years old)^74^ less a number of years expected to retirement item rated by participants (“In approximately how many years do you plan to retire?”). Although intention to retire early might not result in early retirement,^75^ studies have suggested that such intentions predict actual behaviour, hence, adapting intention of early retirement as a useful proxy measure.^76^

### Predictor variables Main Predictor

#### Personal to Perceived Organisational Value Discrepancy in Healthcare Samples (PPOVD-HCS)

In the absence of established measures of the discrepancy between personal and perceived organisational values, the PPOVD-HCS measure was specifically developed. In developing this measure, several steps were taken. First, a taxonomy of healthcare values was derived based on previous literature;^77^ the values listed on websites of international institutions (e.g. WHO, NHS, etc);^78–80^ and NZ-based healthcare websites (e.g. DHBs, New Zealand Medical Association, National Hauora Coalition, etc). Second, additional personal and organisational values linked to compassion in healthcare^24^ were added. Next, this values taxonomy was assessed for face and content validity by a group of senior clinicians, including Indigenous Māori clinicians, to add values seen as important in their work or in organisational functioning. Finally, consolidated healthcare values were compared against universal values from the most commonly used value measure in social sciences – the Portrait Values Questionnaire^81, 82^ – and values from the missing dimensions were added. The last two steps of the process maximised face and content validity, with a list of 23 values ultimately used (see Appendix 3).

In completing this measure, participants rated the personal importance of each value in their work (personal values or PV) before rating how important they thought these same values were to their organisations (perceived organisational values or POV). For both ratings, a 0 to 100 visual analogue scale (VAS) ranging from 0 ‘Not important’ to 100 ‘Extremely important’ were used. An overall discrepancy index was then calculated as a mean of the absolute differences between the 23 perceived organisational and personal value ratings:

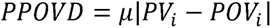

To verify the aggregation of the discrepancy scores, we conducted an Exploratory Factor Analysis using the fa method. The assumptions of normality, linearity, and variables’ being moderately correlated were satisfied. The Kaiser–Meyer–Olkin (KMO) value was 0.97 and Bartlett’s Test of Sphericity was significant (χ2= 18,555.18, df=253, p<0.001), supporting the factorability of the matrix. Screeplot inspection indicated a single component, while parallel analyses revealed two potential components. Interpretatively, a single factor model had better face validity (both models are presented in Appendix 4), explained 54% of the variance (χ2= 2,211.78, df=1025, p<0.001) (Principal Access Factoring method, oblimin rotation were used), and was highly reliable (α = .96) suggesting people who felt value discrepant for one value experienced similar discrepancies for other values.

### Control Variables

#### Socio-demographic data

We collected demographic and occupational data such as gender, age and ethnicity, occupation, years of experience, and work and patient load measured as an average number or hours, or patients seen in a week respectively. Ethnicity data were collected using the NZ population Census Ethnicity question,^83^ with prioritised ethnicity used to assign respondents who report multiple ethnicities to one of either Māori, Pacific, Asian, Middle Eastern Latin American and African (MELAA), ‘Other’ or NZ European (Pākehā) groups (in that order).^84^

#### Organisational variables

Finally, because values and hence, discrepancies, between individual and organisational values might vary as a function of organisational characteristics, we collected data on organisational size (small/medium < 250 employees, large > 250 employees), funding (private or public), setting (community/primary vs. secondary/tertiary), urban vs. rural setting, and belonginess to a cultural framework (e.g. Kaupapa Māori service, Pacific Island Service, Whānau Ora, etc). Belonginess to a cultural framework in the context of this study entails an emphasis on particular set of values (e.g. being in line with Māori or Pacific cultural values, putting emphasis on holistic health, emphasising family values, etc).

#### Social desirability

Because reports of compassion in health samples is prone to desirability bias,^85, 86^ the Marlowe Crowne Social Desirability Scale short form version C (MC-C) was administered. The MC-C is a true or false 13-item measure^87^ in which higher scores indicate a greater tendency towards providing socially desirable responses. The short form has a strong correlation with the full measure (*r* = .91),^88^ good test-retest reliability *r* =.74,^89^ and adequate convergent validity.^90^ Internal reliability estimates tend to be lower in the short form (likely reflecting reductions in content validity), ranging from α = .53 to .67.^85, 86, 88^ Reliability in the present sample was (α = .67).

### Analytic strategy

All analyses and regression assumption checks were performed in R. lm package was used to perform regression analyses. As per our pre-registered analytic plan, outliers were assessed via Tukey fences^91^ and adjusted by ln-transformation; if insufficient, the outliers were capped to 5^th^ and 95^th^ percentiles (i.e. winsorizing).^92^ The variables such as number of patients seen per week and absenteeism were ln-transformed due to skewness and presence of outliers. The variables of absenteeism, weekly hours of work, consideration to retire early, and value discrepancy were additionally windsorized to remove outliers. Missing data were either excluded (i.e. for non-continuous variables or where missing variables represented more than 20% of the data) or imputed via population mean values. If the data were skewed, medians were used instead of means.

Primary analyses proceeded in two main phases. First, we used a combined approach to identify confounds associated with primary and secondary outcomes or the value discrepancy. We selected likely confounds on the outcome variables based on prior work (e.g. gender, years of experience, workload and patient load, and social desirability).^24, 93–102^ Given the likelihood that value discrepancies would vary by ethnicity,^103–105^ we also included ethnicity dummy codes to distinguish between Māori, Pākehā, and Tauiwi (non-Māori/non- Pākehā groups e.g. Pacific, Asian and MELAA, ‘Other’ groupings). We then used univariate two-way t-tests or ANOVAs, and Chi-square analyses to assess possible links between demographic (age, gender), occupational (profession), and organisational (organisational size, setting, funding status and belonginess to a cultural framework) factors and the main predictor.

To test the hypotheses, two-step multiple regressions were performed in R using the lm function. In Step 1, the theoretical and empirical confounders were entered into the model. In Step 2, the main predictor was added to the model, testing whether an overall value discrepancy would predict outcomes and improve model’s fit whilst controlling for other confounders. To ensure scientific rigour, we ran two sets of models – with and without missing data imputation to verify there are no differences in the effects or communicate such differences if there are. Subgroup analyses with regards to participants occupation, ethnicity, and gender were conducted on final models that included predictor variables when there was sufficient statistical power to detect the main predictor effect size (as predicted by the models with the entire sample) for each subgroup. Each analysis was performed separately for each subgroup. Statistical significance was set at PL<L0.05 for all tests. All models were validated by using caret package.

### Deviations from pre-registration

First, although all healthcare organisations were approached at the same time, due to the variation in advertisement timing per institution we recruited more than our minimum sample size to ensure our sample was representative of clinicians across Aotearoa— suggesting that the topic was germane to clinicians.

Second, participants who did not complete the main predictor measure were excluded from the analysis. The reason for this decision reflects elements of the survey flow. To avoid priming, participants only completed the socio-demographic and organisational characteristics data prior to completing the main predictor measure, hence had not completed primary and secondary outcomes. Therefore, anyone who exited the questionnaire prior to completing the main predictors also missed the outcome measures and had to be excluded. Retention analyses indicated no significant differences in demographic or organisational characteristics between clinicians who completed/did not complete the main predictor measure.

Finally, we added an additional secondary outcome of compassion competency as we were interested in whether working in an environment perceived as discrepant affected compassion competency as much as it did compassion ability. We hypothesized that, unlike compassion ability, compassion competency should not be affected by working in a value discrepant environment.

No analyses were conducted prior to these decisions being made. Due to the relatively large sample size we had enough statistical power to predict effect sizes as small as 0.21 with 80% statistical power (95% CI) as assessed via pwr package.

## Results

### Descriptive statistics and univariate analyses

Sample characteristics and descriptive statistics are presented in Table 1. The intervariable correlations table can be found in Appendix 5.

**Table 1:** Sample characteristics and descriptive statistics (N=1025)

|  | **N (%)/**<br>**M(SD) or Median**<br>**(IQR)\*** | **% imputed** |
| --- | --- | --- |
| ***Socio-demographic characteristics*** |  |  |
| Gender |  | 0.0% |
| -male | 154 (15.0%) |  |
| -female | 866 (84.5%) |  |
| -non-binary | 5 (0.5%) |  |
| Ethnicity |  | 0.0% |
| -Pākehā (New Zealand European) | 564 (55.0%) |  |
| -Māori | 156 (15.0%) |  |
| -Tauīwi (non-Māori/non-Pākehā) | 308 (30.0%) |  |
| - Asian | 131 (12%) |  |
| - Other European | 112 (11%) |  |
| - Pacific People | 26 (3%) |  |
| - Middle Eastern, Latin American and African<br>(MELAA) | 18 (2%) |  |
| - Other ethnicity | 21 (2%) |  |
| Age | 44 (13) | 0.1% |
| ***Dispositional measures*** |  |  |
| Social desirability | 21.18 (2.66) | 14.8% |
| ***Occupational characteristics*** |  |  |
| Occupation |  | 0.0% |
| -Doctors | 208 (20.3%) |  |
| -Nurses | 481 (46.9%) |  |
| -Allied and midwives | 336 (32.7%) |  |
| Years of experience | 17.52 (12.76) | 0.2% |
| Patient numbers seen per week* | 20 (12-40) | 0.0% |
| Hours of work per week | 35.92 (8.63) | 0.0% |
| ***Organisational characteristics*** |  |  |
| Organisational size (large >250 vs. small) | 775 (75.6%) | 0.0% |
| Funding type (public vs. private) | 912 (89.0%) | 0.0% |
| Care type (primary vs. secondary) | 339 (33.1%) | 0.0% |
| Urban (vs. rural) | 901 (87.9%) | 0.0% |
| Service belonging to cultural framework | 399 (39.0%) | 0.0% |
| ***Main predictor*** |  |  |
| Experience of value discrepancy | 24.92 (18.68) | 3.0% |
| ***Outcome measures*** |  |  |
| ***Primary outcomes*** |  |  |
| Ability to show compassion | 4.09 (0.58) | 12.8% |
| Burnout | 5.58 (1.69) | 3.3% |
| ***Secondry outcomes*** |  |  |
| Compassion competence | 4.8 (4.4-5.0) | 11.5% |
| Job satisfaction | 42.98 (9.44) | 3.2% |
| Absenteeism (in days)* | 7 (3-11) | 0.0% |
| Intention to retire early (in years) | 0.14 (4.35) | 0.5% |

Proportion Z tests and t-tests indicated that our sample of clinicians had on average more female doctors (61% vs. 47%, χ^2^(1)= 18.58, p < 0.001), slightly younger doctors ((M = 46, μ=43, SD=13), t(1024) = 7.4, p < .001, Cohen’s D = 0.2.), and more Māori doctors and nurses (12% vs. 7%, χ^2^(1)= 1067.7, p < 0.001) then the respective populations ^106, 107^. No national statistics are currently collected about allied health professionals in NZ to provide a comparison.

Assessing whether PPOVD scores covaried with demographic and occupational variables, ANOVAs yielded significant mean differences in value discrepancies by gender (*F*(1,1018)=4.5, P=0.03), ethnicity (*F*(1,1022)=7.9, P<0.001), occupation (*F*(2,1022)=3.2, p=0.04), organisation size (*F*(1,1023)=16.2, P<0.001), organisational funding (*F*(1,1022)=10.4 P=0.001), and organisational belonginess to cultural framework (*F*(1,1020)=33.4, P<0.001). No mean differences were observed between healthcare professionals working in urban versus rural environment (P=0.36) or whether they worked in primary, secondary or tertiary care setting (P=0.66). Based on these analyses, occupation, organisational size, funding status, and belonginess to cultural framework were added as confounds to the primary regressions.

### Regression results

#### H1A: Does working in a value discrepant environment predict lower perceived ability to provide compassion?

At Step 1 (see Table 2A), the regression model with confounders explained 14.04% of the variance of healthcare professional ability to show compassion, *F*(12,1003) = 14.81, p < 0.001. Adding the value discrepancy score at Step 2, the model explained 18.33% of the variance in the ability to show compassion, *F*(13,1002) = 18.53, P<0.001, a significant improvement in model fit, *R*2Δ = 4.29%, *F*Δ(1,1002) = 53.75, P< 0.001). As hypothesized, a greater discrepancy between individual and organisational values predicted lower reported ability to show compassion even after controlling for a range of confounds. Cohen’s f2 effect size for value discrepancy was .05.

**Table 2A.**
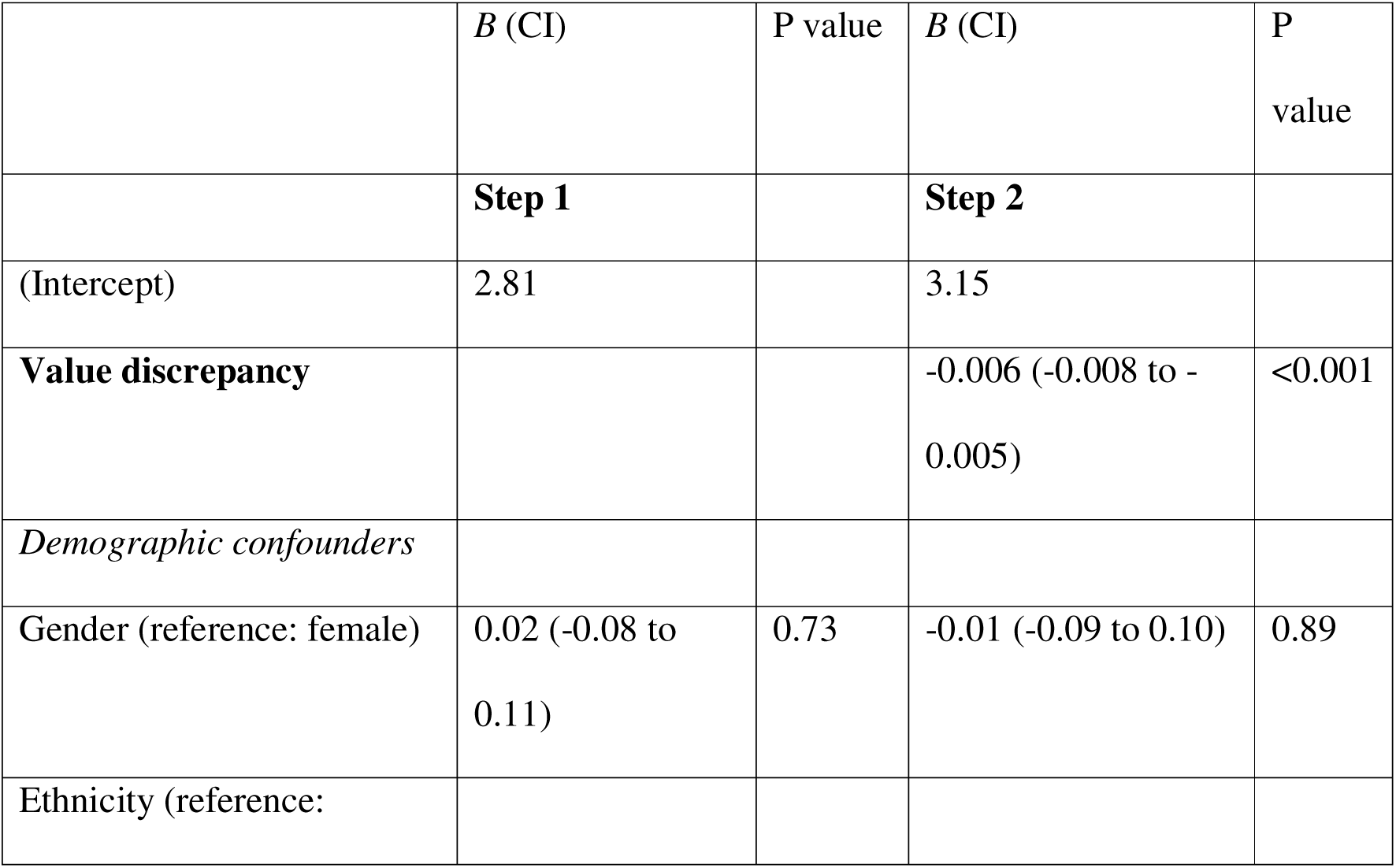
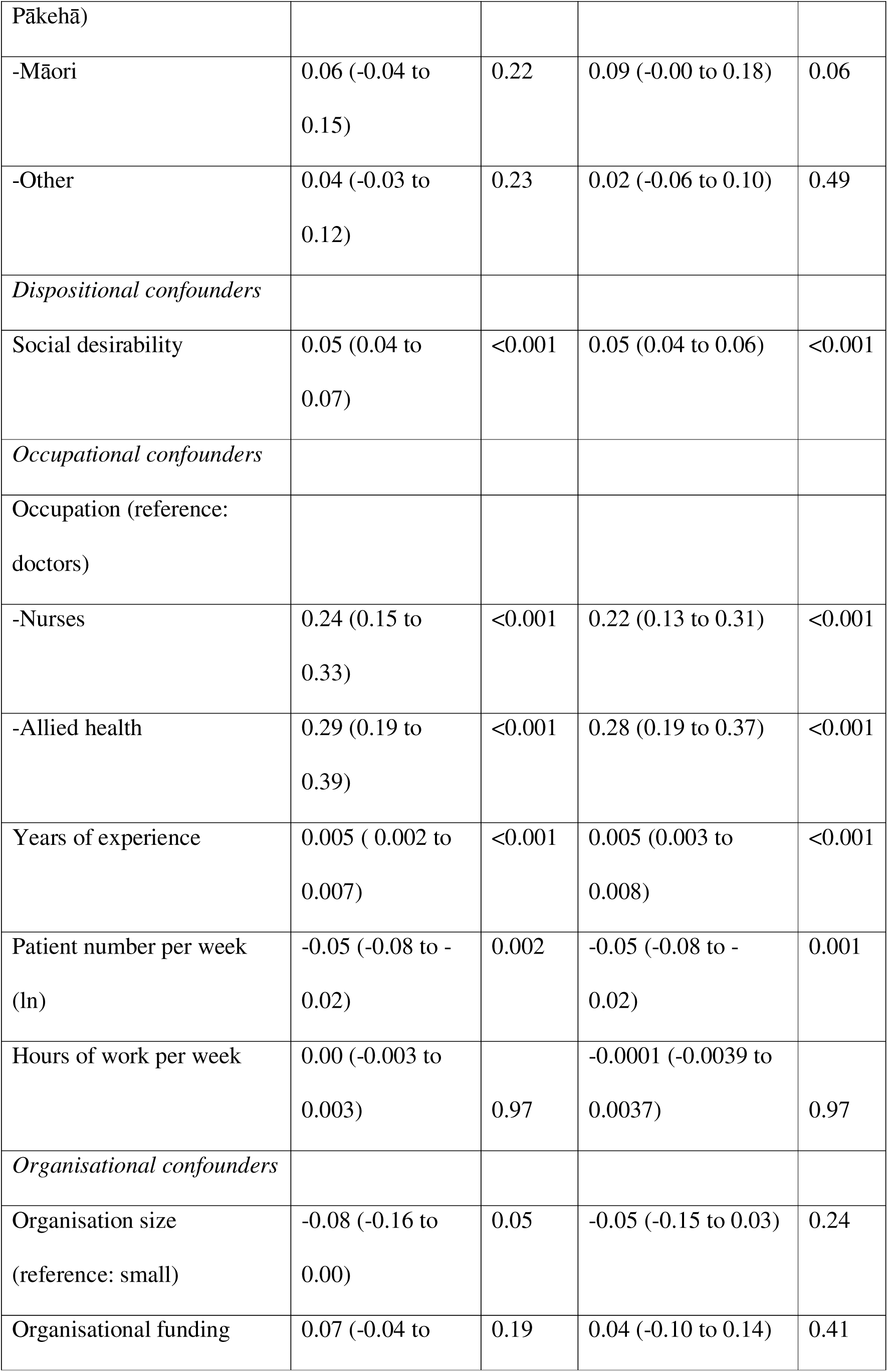
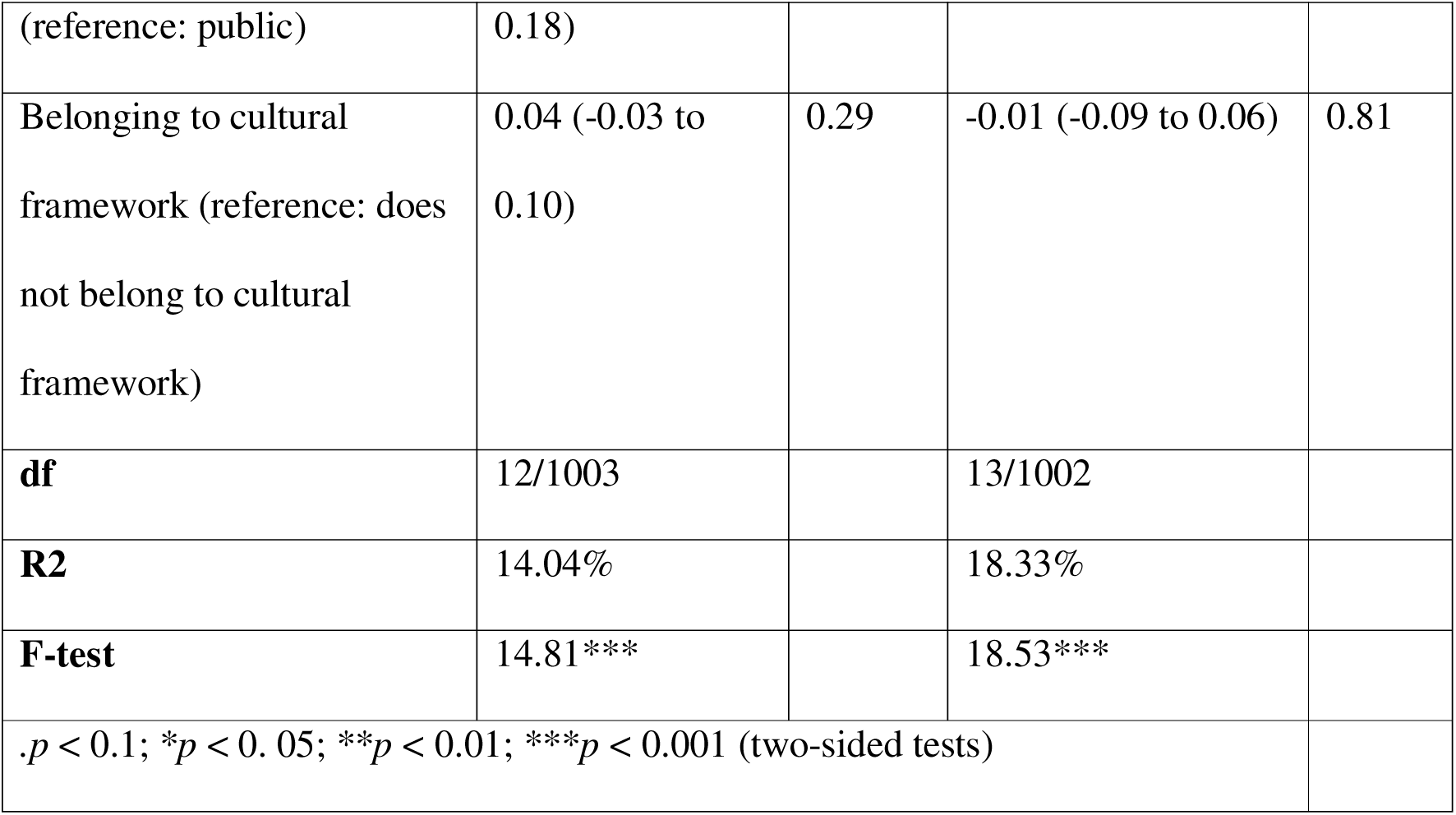
Effect of value discrepancy on the ability to provide compassion

#### H1B: Does working in a value discrepant environment predict lower compassion competence?

At Step 1, the regression model with confounders explained 16.32% of the variance of healthcare professional compassion competence, *F*(12,1003) = 17.49, p < 0.001. Adding the value discrepancy score at Step 2, the model explained 16.34% of the variance in compassion competency, *F*(13,1002) = 16.25, P<0.001, showing no improvement in model’s fit, *R*2Δ = 0.02%, *F*Δ(1,1002) =1.24, P=0.25). As hypothesized, the discrepancy between individual and organisational values did not predict a difference in compassion competency (see Table 2B).

**Table 2B.**
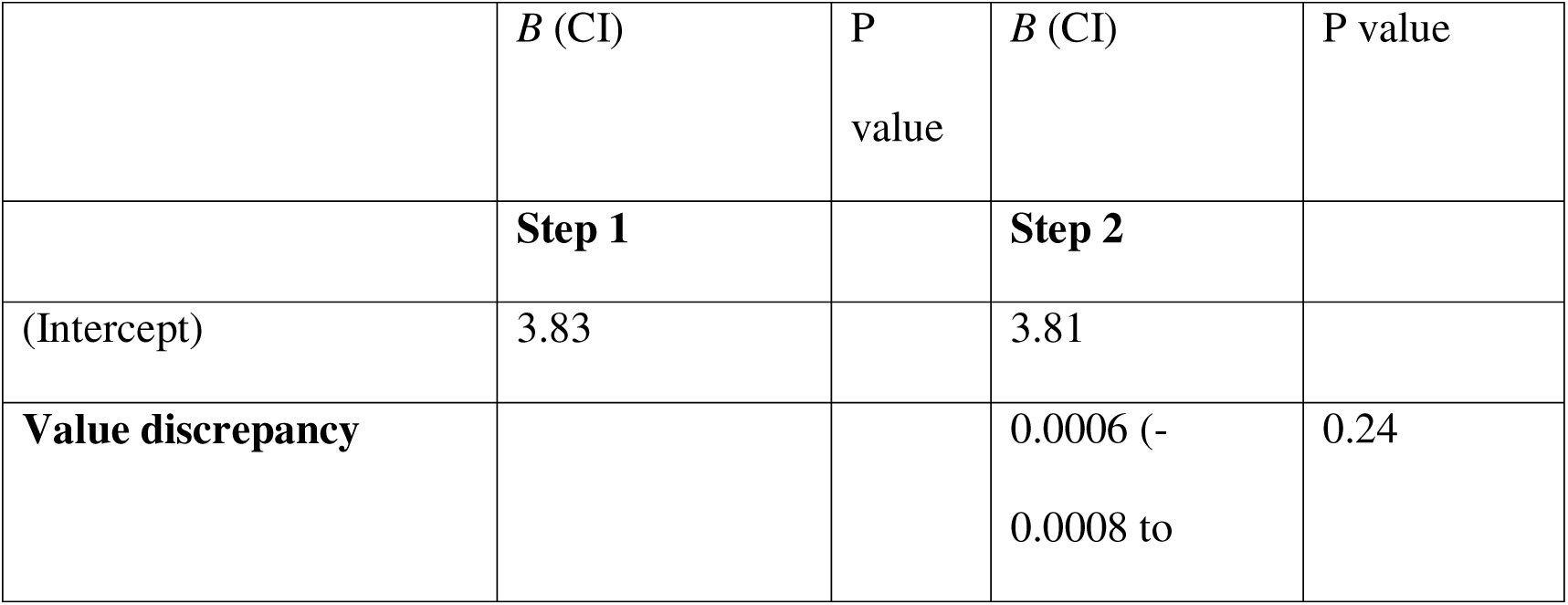
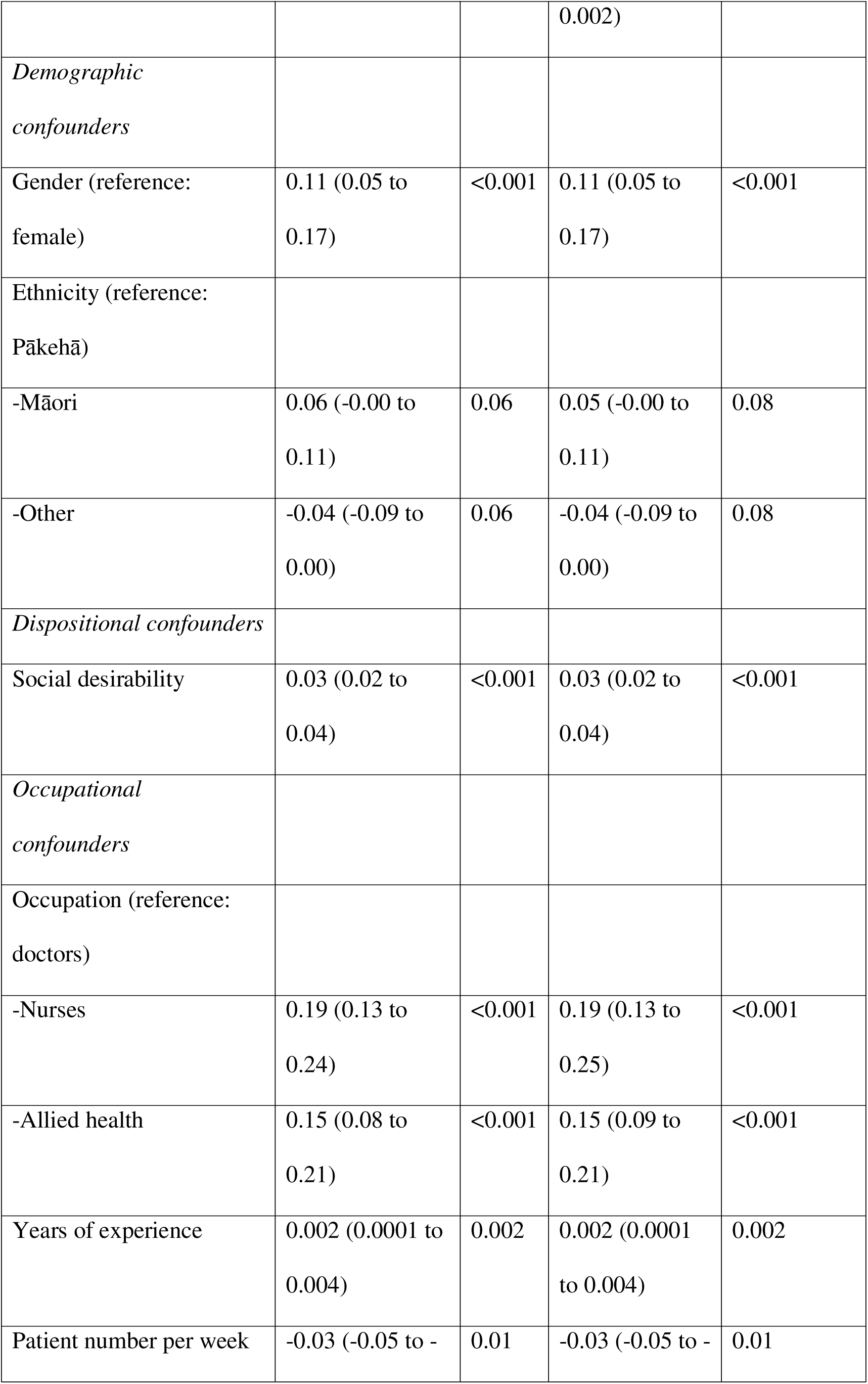
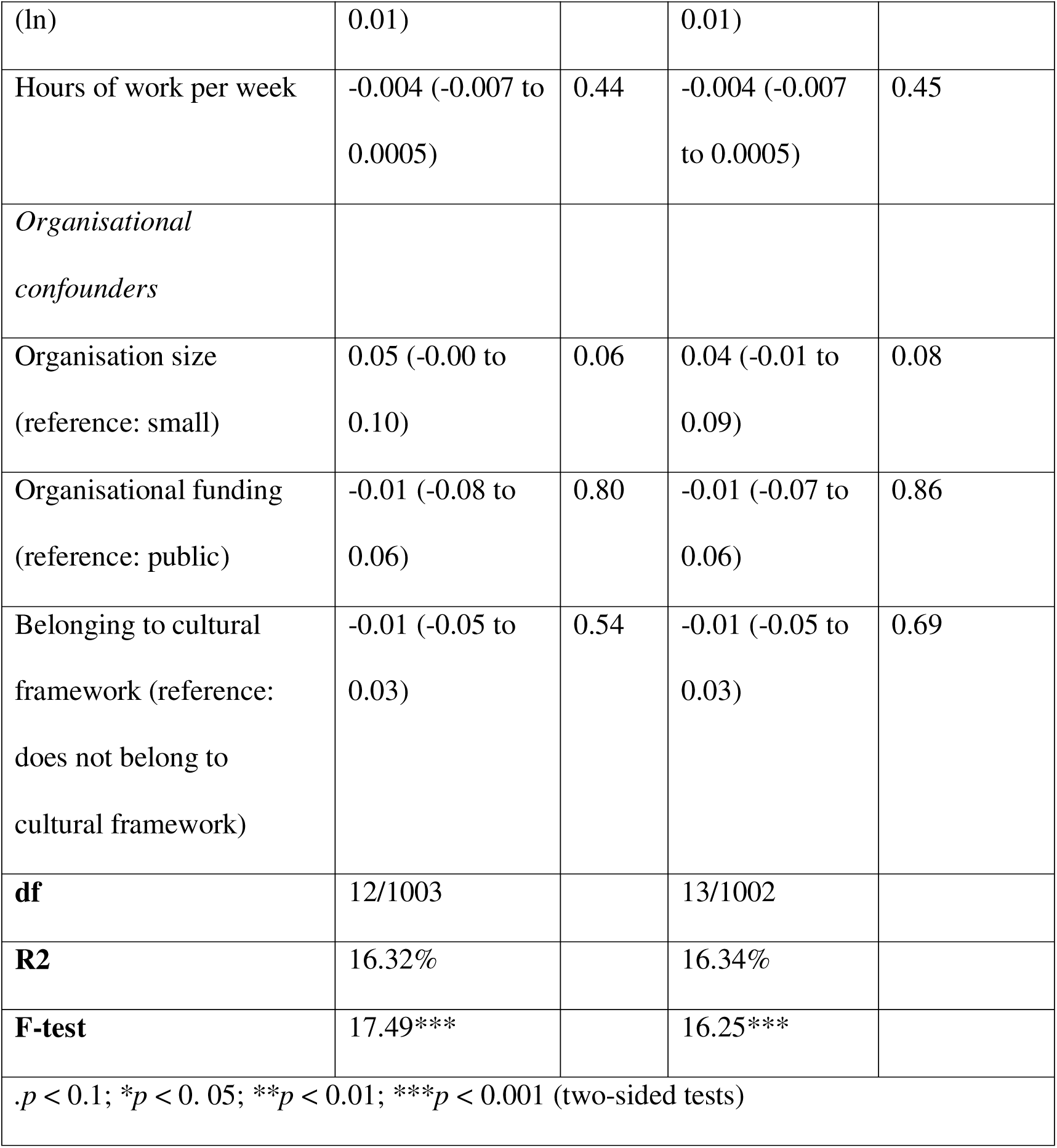
Effect of value discrepancy on compassion competence

#### H2: Does working in a value discrepant environment predict burnout?

At Step 1 (see Table 2C), the regression model explained 13.09% of the variance in burnout, *F*(12,1003) = 13.74, P< 0.001. Adding value discrepancy measure in Step 2, the model explained 18.06% of the variance, *F*(13,851) = 16.06, P<0.001, an improvement in fit (*R*2Δ = 4.97%, *F*Δ(1,1002) = 61.77, P < 0.001). As hypothesized, experiencing a greater discrepancy between personal and organisational values was associated with greater burnout, even after controlling for multiple confounds. Cohen’s f2 effect size for value discrepancy was .06.

**Table 2C.**
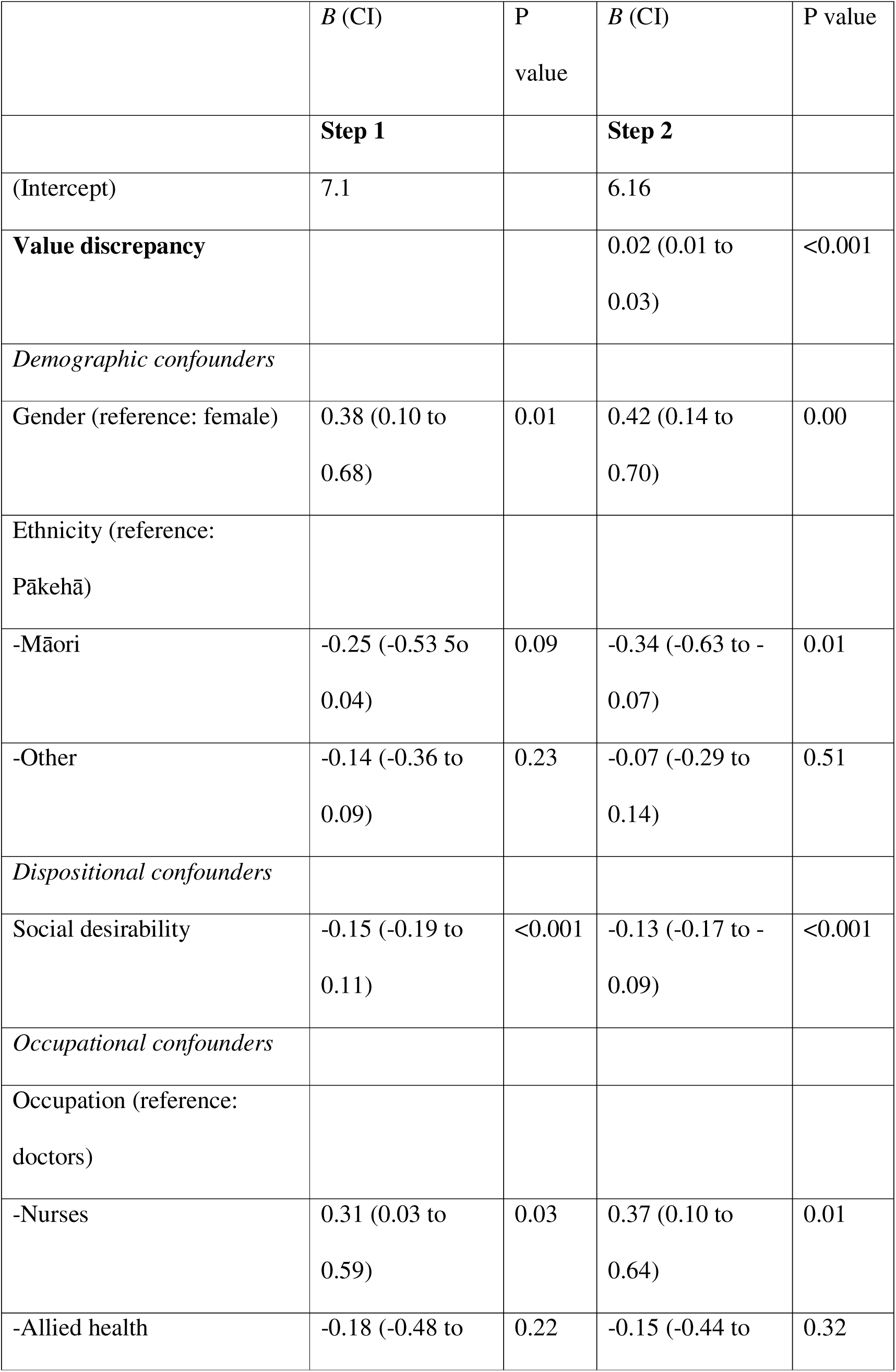
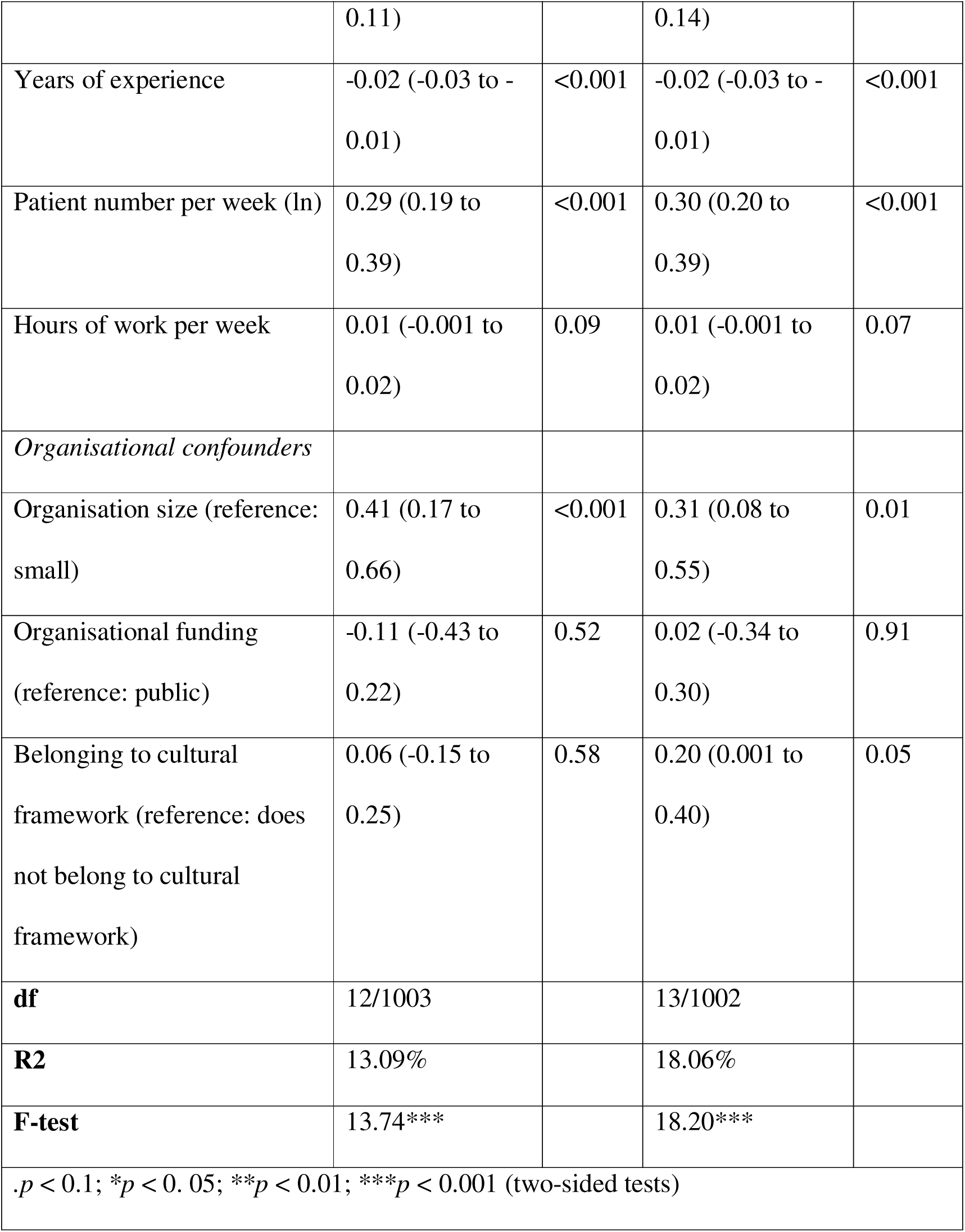
Effect of value discrepancy on professional burnout

#### H3: Does working in a value discrepant environment predict job satisfaction?

At Step 1 (see Table 2D), the regression model explained only 3.50% of the variance of healthcare professionals job satisfaction, *F*(12,1003) = 4.04, P < 0.001. Adding value discrepancies in Step 2, the model explained 16.93% of the variance*, F*(13,1003) = 16.92%, P<0.001; the model fit was improved (*R*2Δ = 13.42%, *F*Δ(1,1002) = 163.53, P<0.001). As hypothesized, greater value discrepancies predicted lower job satisfaction even after controlling for confounds. Cohen’s f2 effect size for value discrepancy measure was .14.

**Table 2D.**
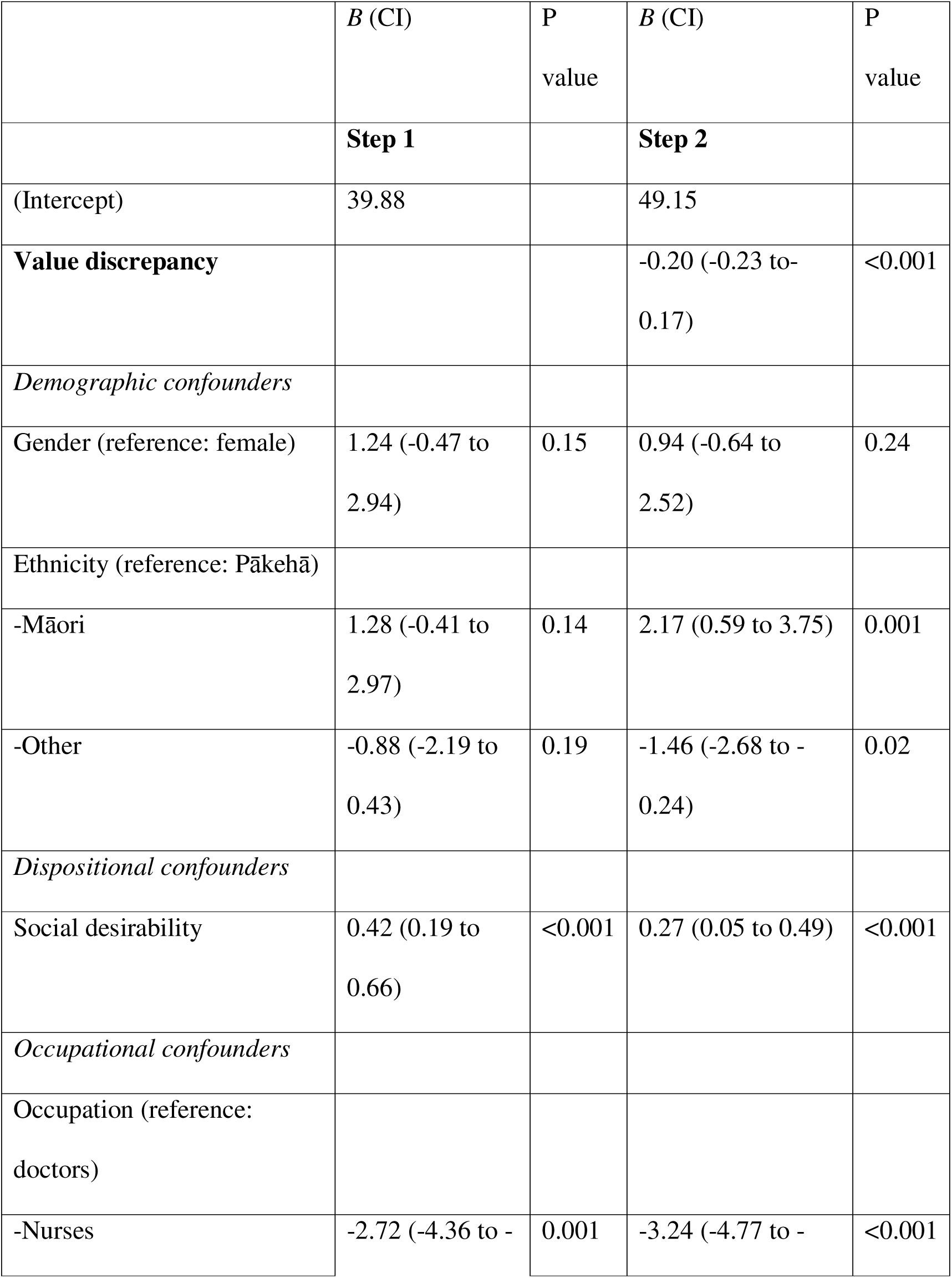
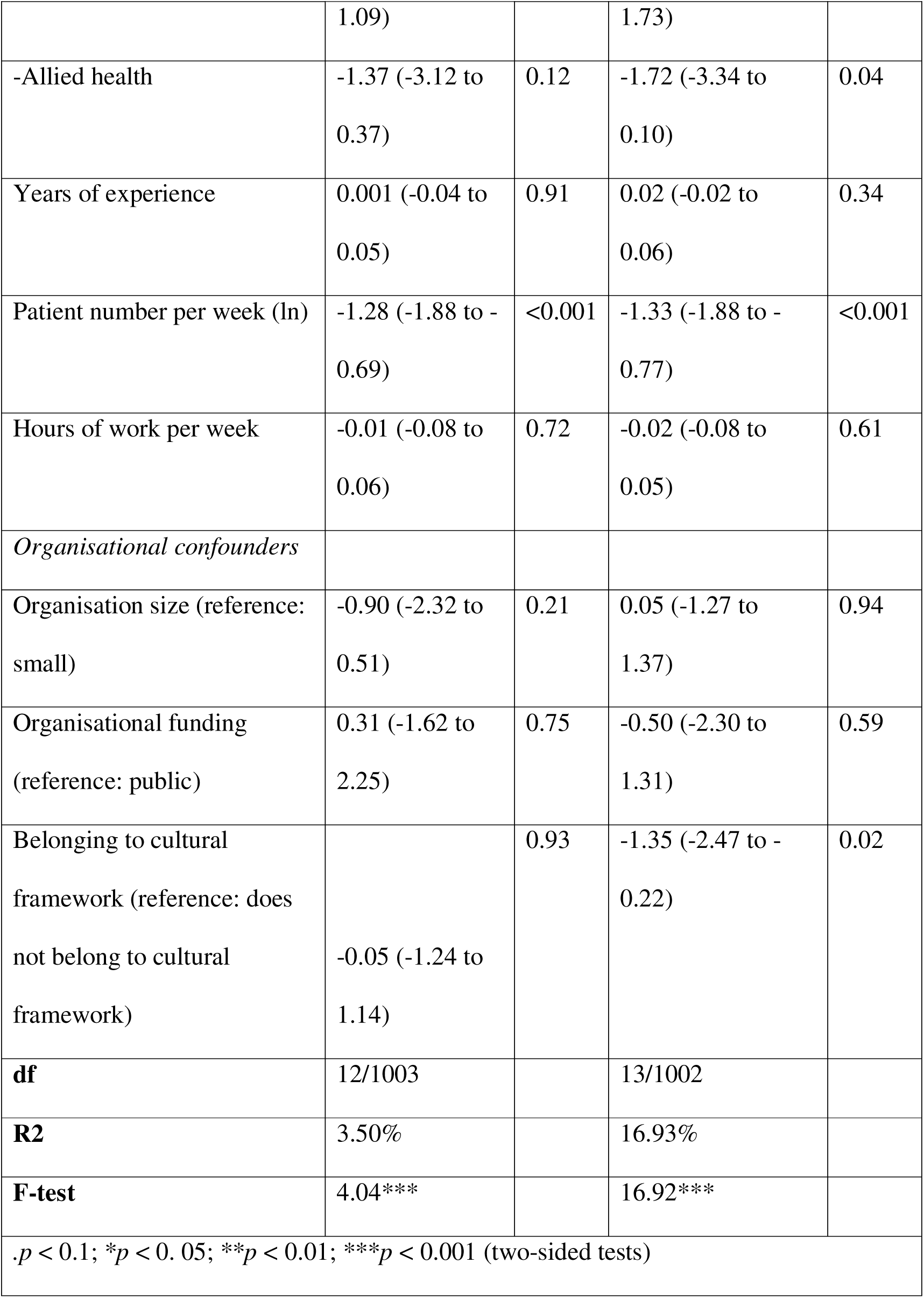
Effect of value discrepancy on job satisfaction

#### H4: Does working in a value discrepant environment predict absenteeism?

At Step 1 (see Table 2E), the regression model explained 14.63% of the variance of healthcare professionals absenteeism, *F*(12,1003) = 15.50, P < 0.001. Adding value discrepancy variable at Step 2, the model explained 15.08% of the variance, (*F*(13,1002) = 14.86, P<0.001), with slight improvement in model’s goodness of fit (*R*2Δ = 0.45%, *F*Δ(1,1002) = 6.27, P=0.01). Value discrepancy predicted higher absenteeism after controlling for other confounders. Cohen’s f2 effect size for value discrepancy was negligible .01.

**Table 2E.**
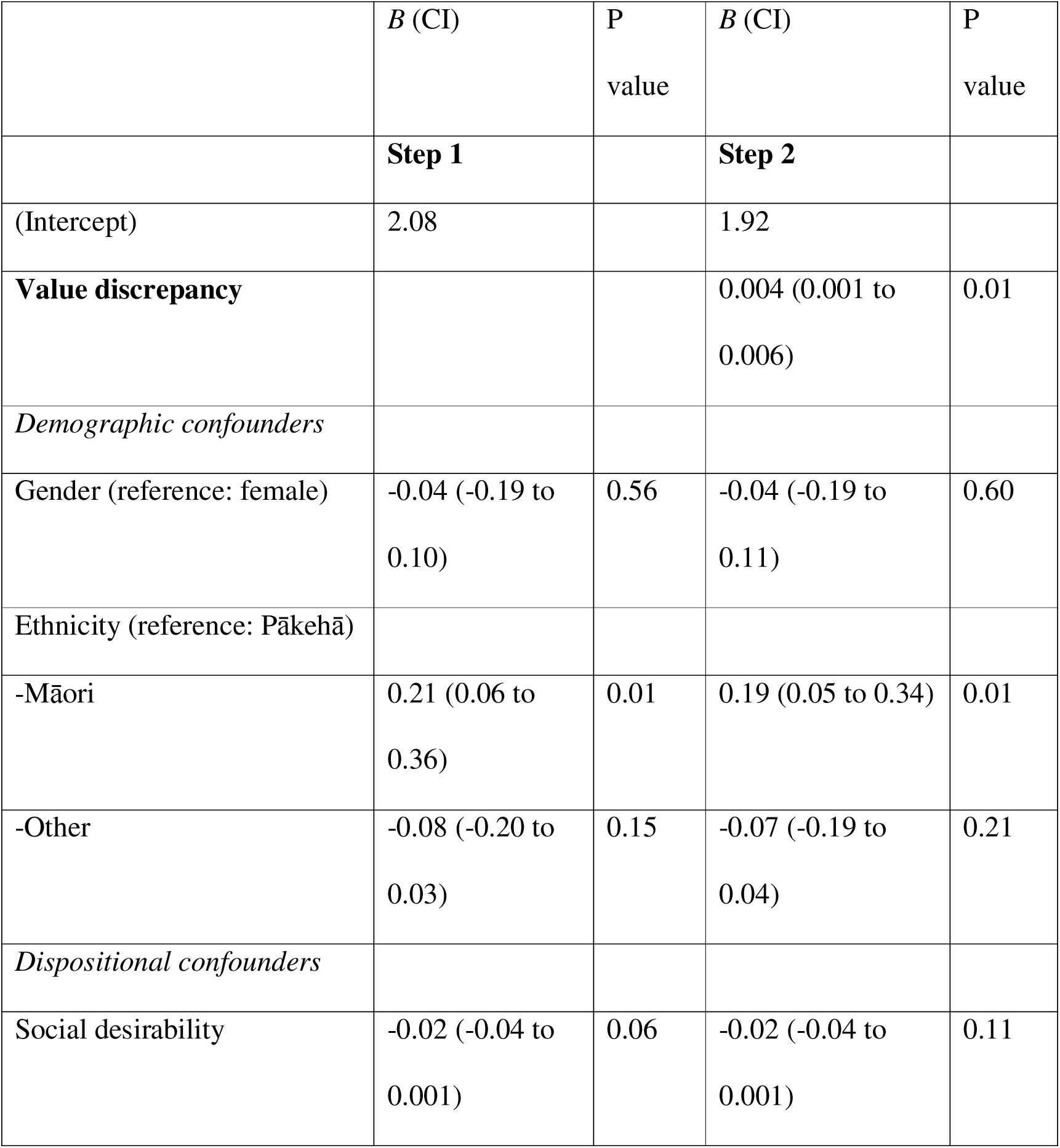
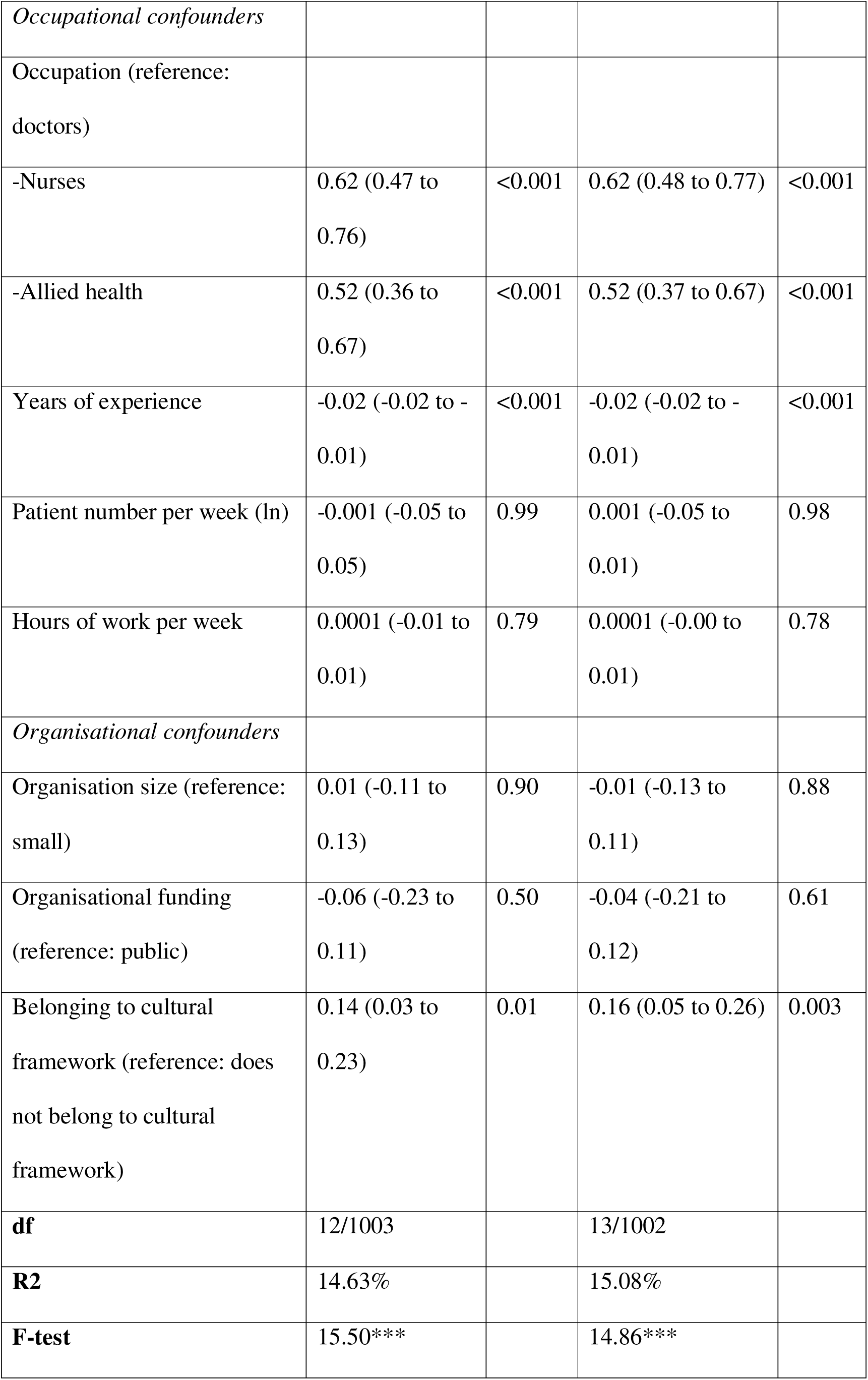
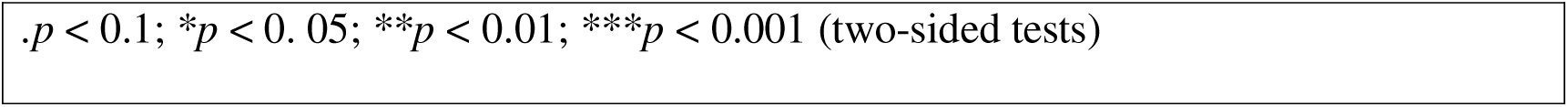
Effect of value discrepancy on absenteeism (ln)

|  | *B* (CI) | P value | *B* (CI) | P value |
| --- | --- | --- | --- | --- |
|  | **Step 1** |  | **Step 2** |  |
| (Intercept) | 2.08 |  | 1.92 |  |
| **Value discrepancy** |  |  | 0.004 (0.001 to 0.006) | 0.01 |
| *Demographic confounders* |  |  |  |  |
| Gender (reference: female) | -0.04 (-0.19 to 0.10) | 0.56 | -0.04 (-0.19 to 0.11) | 0.60 |
| Ethnicity (reference: Pākehā) |  |  |  |  |
| -Māori | 0.21 (0.06 to 0.36) | 0.01 | 0.19 (0.05 to 0.34) | 0.01 |
| -Other | -0.08 (-0.20 to 0.03) | 0.15 | -0.07 (-0.19 to 0.04) | 0.21 |
| *Dispositional confounders* |  |  |  |  |
| Social desirability | -0.02 (-0.04 to 0.001) | 0.06 | -0.02 (-0.04 to 0.001) | 0.11 |
| *Occupational confounders* |  |  |  |  |
| Occupation (reference: doctors) |  |  |  |  |
| -Nurses | 0.62 (0.47 to 0.76) | <0.001 | 0.62 (0.48 to 0.77) | <0.001 |
| -Allied health | 0.52 (0.36 to 0.67) | <0.001 | 0.52 (0.37 to 0.67) | <0.001 |
| Years of experience | -0.02 (-0.02 to -0.01) | <0.001 | -0.02 (-0.02 to -0.01) | <0.001 |
| Patient number per week (ln) | -0.001 (-0.05 to 0.05) | 0.99 | 0.001 (-0.05 to 0.01) | 0.98 |
| Hours of work per week | 0.0001 (-0.01 to 0.01) | 0.79 | 0.0001 (-0.00 to 0.01) | 0.78 |
| *Organisational confounders* |  |  |  |  |
| Organisation size (reference: small) | 0.01 (-0.11 to 0.13) | 0.90 | -0.01 (-0.13 to 0.11) | 0.88 |
| Organisational funding (reference: public) | -0.06 (-0.23 to 0.11) | 0.50 | -0.04 (-0.21 to 0.12) | 0.61 |
| Belonging to cultural framework (reference: does not belong to cultural framework) | 0.14 (0.03 to 0.23) | 0.01 | 0.16 (0.05 to 0.26) | 0.003 |
| **df** | 12/1003 |  | 13/1002 |  |
| **R<sup>2</sup>** | 14.63% |  | 15.08% |  |
| **F-test** | 15.50*** |  | 14.86*** |  |

. $p < 0.1$ ; \* $p < 0.05$ ; \*\* $p < 0.01$ ; \*\*\* $p < 0.001$ (two-sided tests)

#### H5: Does working in a value discrepant environment predict intention to retire early?

At Step 1 (see Table 2F), the regression model explained 4.77% of the variance of healthcare professionals absenteeism, *F*(12,1003) = 5.24, P < 0.001. Adding the value discrepancy variable at Step 2, the model explained 5.10% of the variance, *F*(13,1003) = 5.19, p<0.001, an improvement in model fit (*R*2Δ = 0.33%, *F*Δ(1,1002) = 4.31, P=0.04). Greater value discrepancy measure predicted greater intention to retire early while controlling for other confounders. Cohen’s f2 effect size for value discrepancy was negligible .004.

**Table 2f.**
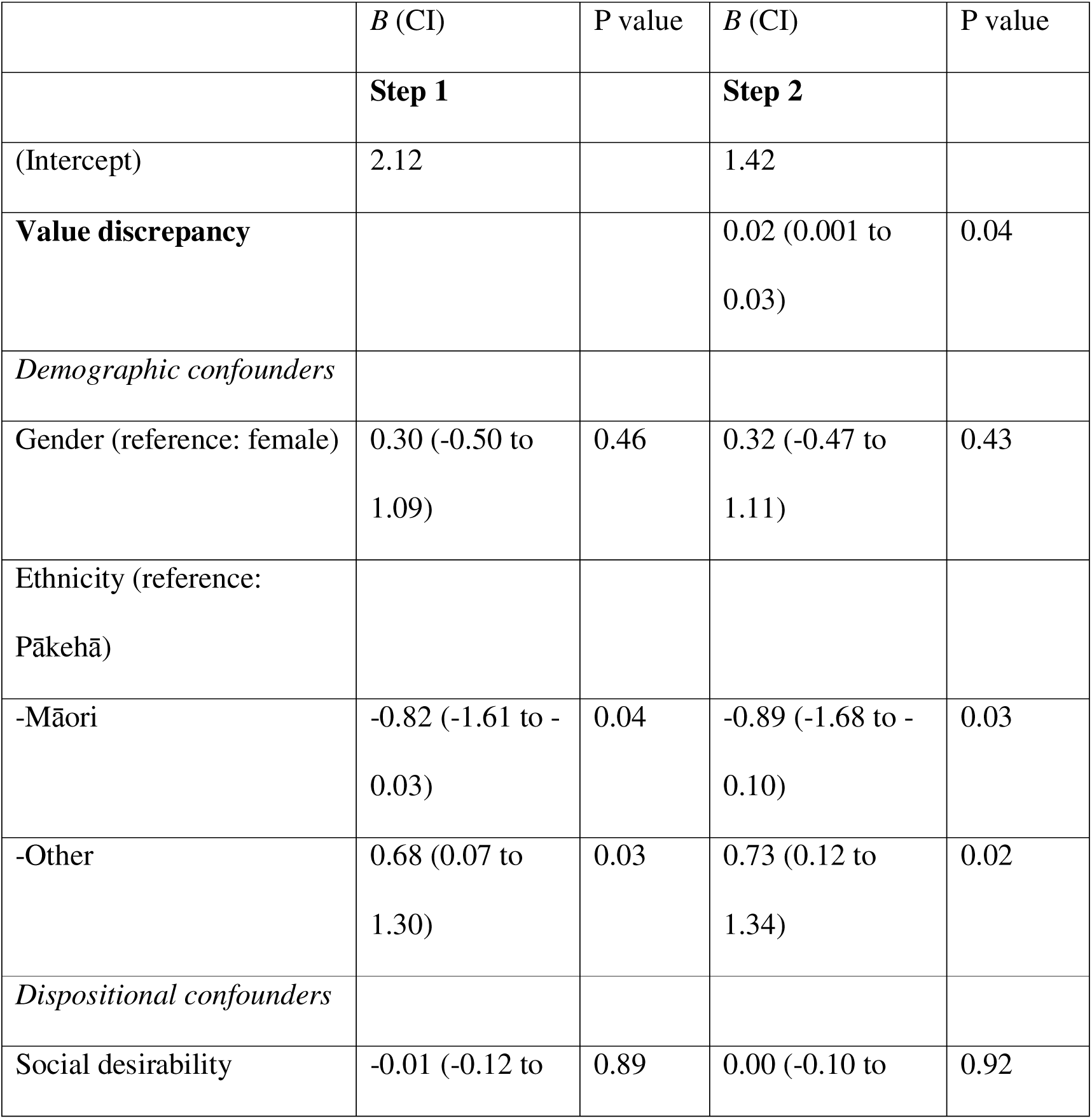
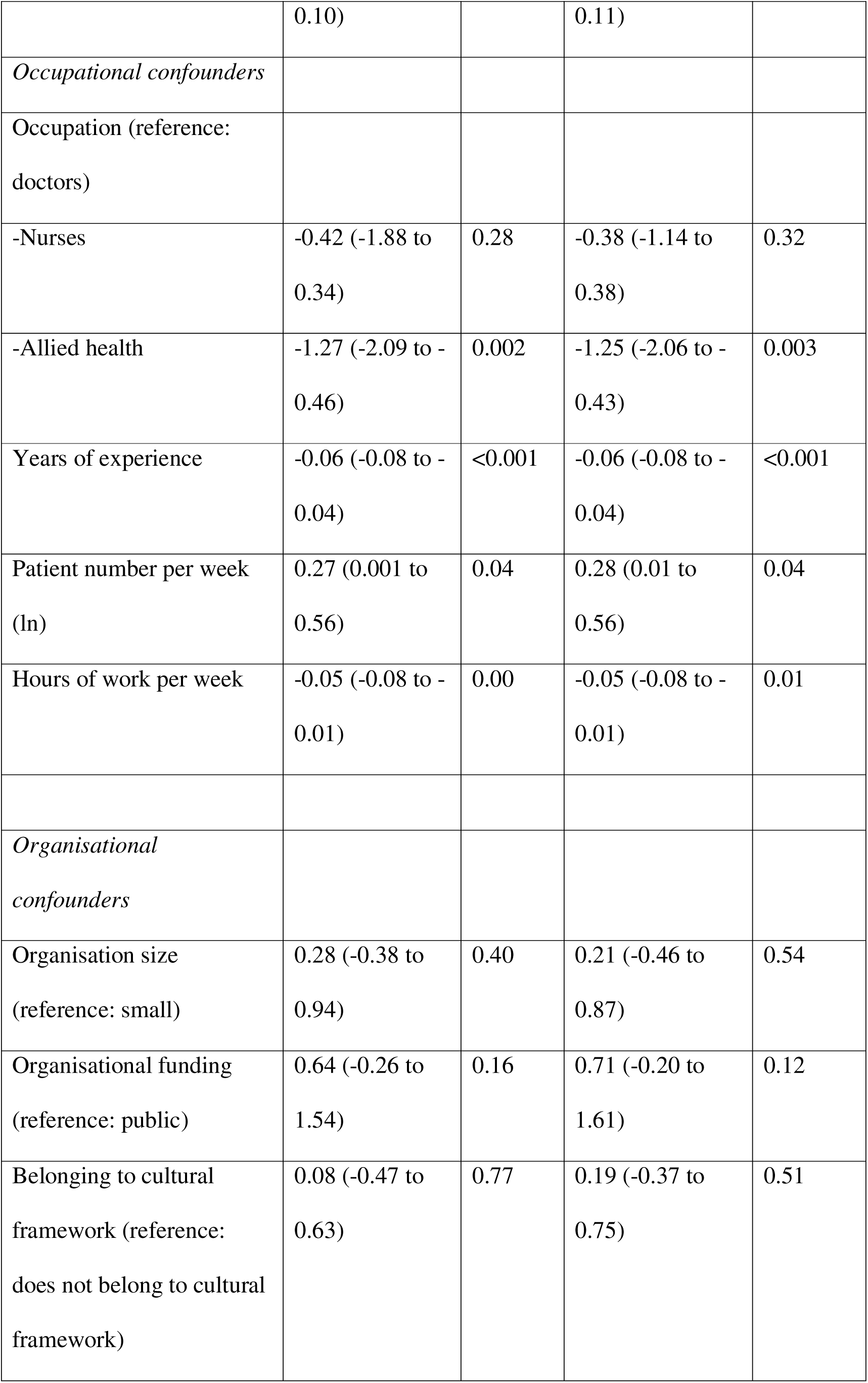
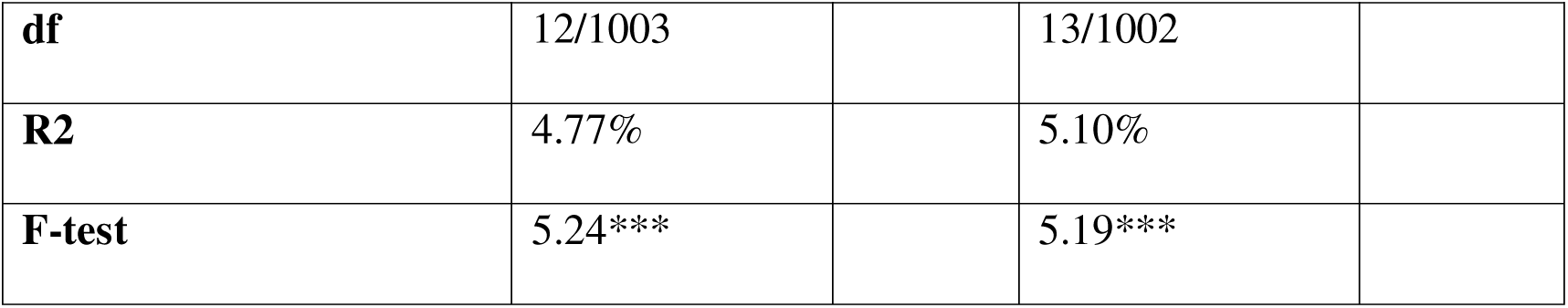
Effect of value discrepancy on early retirement

The models with and without missing data imputation produced similar results. Subgroup analyses for models related to the outcomes of compassion ability, compassion competence, burnout, and job satisfaction conducted per each subgroup of occupation, ethnicity, and gender were consistent with those in the overall sample.

## Discussion

Prior work has implied that the inability to act in accordance with one’s values creates a ‘stress of conscience’ or moral distress, and contributes to clinician burnout.^22, 50–52^ However, clear empirical tests operationalising this possibility have been lacking. Thus, an important contribution of the current study lies in providing a robust empirical demonstration that working in healthcare environments experienced as discrepant with personal values has negative effects on key outcomes including the ability to provide compassionate medical care, job satisfaction, burnout, absenteeism, and early retirement intention. In terms of extending the rigour with which such questions are tested, these effects held in a large sample even after controlling for a host of demographic and professional confounds, including social desirability.

While the exact reasons that working in value discrepant healthcare environments predicts poorer outcomes for clinicians are not yet clear, our sense is that there are two interrelated possibilities – one reflecting the psychological stress that value discrepancies create and one reflecting lowered motivation and satisfaction with the core tasks and contingencies the working environment entails. Below, we consider these possibilities more fully in light of prior theory and data in order to further our understanding of how work environments impact compassion specifically and clinician wellbeing more broadly as well as in terms of what this might imply regarding potential interventions.

The notion that engaging in behaviours that are inconsistent with beliefs/values is psychologically costly is well-established in research.^49^ Theories such as the Conservation of Resources (COR) theory^108^ suggest that operating in environments that produce tensions of this kind require the expending of cognitive and emotional resources. Meta-analytic studies report that chronic exposure to conflicting work demands is associated with psychological resource depletion and, consequently, emotional and cognitive fatigue^109, 110^ that likely contribute to poorer outcomes, as was observed in the current study.

Equally, it is important to remember that the odds that particular behaviours (such as compassionate behaviours) occur are partly determined by the expectation that that behaviour will lead to desired (positive) outcomes.^111, 112^ Organisational values are critical to such processes insofar as such values are reflected in the contingencies – performance and safety targets, task allocations and priorities, reward and recognition criteria etc – that dominate the professional landscape. Our results indicate that clinicians’ subjective competency in providing compassionate care may remain high. However, when working in value-discrepant environments, the employment contingencies we describe above may require that clinicians repeatedly compromise personal values and refrain from using available competencies in the service of keeping performance in line with requirements. As noted, operating in this way over long periods of time requires ongoing psychological regulation with the discrepancies between one’s own values and those embedded in the professional environment. Prior studies suggest that perceptions of low personal control predict distress^22^ and poorer wellbeing,^113^ likely creating strain and contributing to outcomes such as burnout^114^ and an inability to express compassion.^22^ In turn, such effects likely contribute to more systemic problems such as productivity loss, employee turnover, and an increase in medical errors and malpractice claims that, albeit more indirectly, result in greater healthcare expenditures and further reduce patient access to timely care.^8, 19, 114–117, 118^

Moreover, in situations when one’s resources (notwithstanding whether these are psychological, social, or material), are threatened or gains after investing effort are minimal, maladaptive coping becomes more likely ^109^. Research in medical compassion ^24, 26, 97^ shows that self-protective strategies like avoidance and detachment are common in healthcare samples ^119^, impair compassion ^120–125^ and predict burnout ^126^. As such, both psychological tension and low expectations regarding positive outcomes seem likely to contribute to the link between being situated in value-discrepant environments and poorer professional outcomes.

Importantly, none of the explanations offered here (nor indeed the findings themselves) are consistent with the notion of compassion fatigue as reflecting the cost of caring that arises from exposure to repeated suffering.^21, 127^ Indeed, prior work shows that expressing compassion in healthcare facilitates more compassion^128, 129^ and that prosocial dispositions may prevent burnout^130–132^ suggesting that providing compassion does not deplete healthcare professionals but sustains them. Our suspicion here is that what is impeding compassion in healthcare is compassion distress^22^ – the inability to provide compassion due to it being obstructed by situational factors. Future studies directly testing this possibility would be a fertile area of research, helping to challenge discourse that has led to individualised rather than systemic approaches to addressing problems of lack of compassion and impaired professional wellbeing in healthcare samples.^133^ Additionally, our results suggest that improving compassion in healthcare is not simply dependent on motivating individual health professionals to be more (self)-compassionate or teaching rote compassion skills but, rather, creating the organizational and clinical conditions where compassion can flourish.^23, 86, 134, 135^ Our empirical study represents the first known results that attempt to bridge the gap between the personal and environmental effects on compassion and other key healthcare professionals’ outcomes.

### Implications for practice

Our findings suggest that addressing the incongruence between personal and perceived organisational values could be of considerable benefit to healthcare organisations. The current climate of austerity and uncertainty^43^ combined with the additional service demands created by COVID-19 pandemic^44^ suggest that healthcare organisations will be striving for even greater efficiency and cost savings. Discrepancies in values are likely to be exacerbated by variations in resource allocation^109^ and increasing resource scarcity.^44^ In fact, this dynamic was already clearly seen in evidence during the rationalisation of health care resourcing that occurred during COVID-19 and by making decisions about who gets health care during COVID-19, in what ways, and how it would affect clinicians.^136–141^ Unfortunately, again, the value set informing those discussions and the resultant algorithms have shown to be more ‘business as usual’, process-orientated, tokenistic and inequitable, likely worsening the pre-existing tensions between organisational values and those of the healthcare workforce. Given that that the experience of values discrepancies can have significant costs for clinicians, patients and, indirectly, for healthcare systems, these dynamics must be addressed.

Although there is little prior work in this area, we suspect that the most effective way to positively affect patients’ and clinicians’ wellbeing and reduce the costs associated with values discrepancies will be for healthcare organisations to both revisit their values in light of these data and, more importantly, systematically assess and report whether their stipulated values are reflected in policy, targets, and priorities throughout the system. Put another way, rather than focusing on top-down rewriting of mission statements, job boards, and email signatures that *already* signal humanistic values,^39–42^ healthcare organisations need to invest in a careful examination of whether and how these values are *embedded* and operationalized in the day-to-day practices, behaviours, beliefs, decisional processes, and core performance indices^142^ including, patient outcomes. Consequently, a recommended starting point to begin to tackle this complex problem is to bring the internal healthcare organisations’ priorities into line with humanistic values that are also sought after by staff. We argue that one way to do so is to measure the associations between value alignment and outcomes. In addition, some authors also suggest that adaptation of certain organisational practices such as, value-based feedback^143^ might be helpful in fostering compassion and aligning it with other suggested measures. Finally, healthcare organisations need to start supplementing the predominant economic values assessments^144–146^ by systematically measuring and valuing compassion both from the perspective of clinicians and the patients they serve.

In a broader sense, our data also suggest that policymakers and government agencies should reconsider how policy influences priorities as well as targets embedded in funding criteria within healthcare organisations. External pressures, notably those of a fiscal nature, profoundly impact how organisations operate, leading to shifts in organisational vision, values, and goals.^46, 147, 148^ However, as external pressures mount, healthcare organisations are likely to continue to prioritise fiscal considerations at the expense of patient and healthcare provider wellbeing—creating environments and values that are out of step with the values of patients and healthcare professionals. Given the evidence presented here that discrepancies may contribute to burnout and a lower ability to provide compassion to patients, the benefit of short-term austerity solutions may not outweigh the long-term costs to patients and healthcare professionals. Due to the neoliberal reforms that occurred in the 1980s and 1990s healthcare in NZ became predominantly viewed through an economic lens and in terms of productivity—requiring broader fiscal reforms in the future.^146, 149, 150^ However, as research has repeatedly indicated, measures of organisational efficiency do not necessarily lead to improved outcomes.^24, 114^ Our study confirms that serving a greater number of patients, although an important performance target, might actually compromise clinicians’ wellbeing and their ability to be compassionate. In viewing healthcare as a form of industrial production we are potentially setting up a system (and the people it serves) for failure. If compassion truly is paramount, this needs to be identified as a priority not simply in the mission statement but also in the budgets and the associated performance indices.

We have previously argued that the lack of compassion in healthcare is a systemic problem requiring systemic solutions.^133, 151^ The same appears to be true with respect to the effects of value discrepancies. For example, while individualised interventions such as increased resilience might be useful in managing environments in which personal and organisational values diverge, evidence suggests staff may struggle to implement sustainable changes where organisational environments are unsupportive (or compete) with such changes.^152^ This study echoes the results of a recent review that has shown that compassion training will be optimized when it “engages institutional participation, improves leadership at all levels, adopts a multimodal approach, and uses valid measures to assess outcomes”.^153^ Hence, focusing on systemic rather than individual-level interventions may be a more effective, scalable, and cost-efficient way to deal with the consequences of values discrepancies, clinician burnout and to ultimately improve compassion to patients.

### Limitations

Although this is the first known empirical study investigating how working in value discrepant environments impact compassion and professional wellbeing, this study is not without limitations. First, although our large sample was recruited from all health districts in NZ, it is nonetheless self-selected, differentially omitting those too busy, burnt out, or cynical to participate. We also note that the recruitment took place during the Covid-19 Delta outbreak in NZ which might have differentially impacted the recruitment of Māori and Pacific respondents, many of whom stood up to the challenge to provide care for their respective communities who were disproportionately affected by the pandemic.^137, 154^ However, the study was designed to mitigate these risks by having a 3-month long recruitment window with multiple follow ups and recruiting via healthcare and professional organisations as well as via unions and alumni lists. By advertising the study as involving ‘Institutional barriers to care for kaimahi hauora (healthcare workers) in Aotearoa’ and not explicitly mentioning compassion, we tried to avoid acquiring a heavily prosocial sample. Nonetheless, advertising the study in this way may have differentially recruited professionals who were less satisfied with employers. The inclusion of previously-tested recruitment strategies such as provision of a prize draw and inclusion of some te reo Māori (the Māori language) in our study advertisements was important for reaching Māori health professionals.^155, 156^

We also acknowledge that the results of this survey reflect the NZ population with its specific culture and the ways in which the predominantly public healthcare system operates.^146^ However, while our findings might not be generalizable to other contexts and health systems, the universal importance of humanistic values at personal as well as healthcare organisational levels^24^ and the comparable external and fiscal pressures that are imposed on healthcare systems worldwide may suggest similar patterns are likely elsewhere. Nonetheless, future studies should replicate this approach in other countries and jurisdictions for cross-cultural comparison.

Additionally, while the cross-sectional, self-report design allowed for the rapid acquisition of a large sample (and social desirability was controlled), the data remain fundamentally correlational precluding certainty about causality. Moreover, although the effect of value discrepancies on the key performance indicators such as absenteeism and intention of early retirement were significant, the size of effect was negligible. Future research should explore whether the effect of value discrepancies on these metrics are mediated by other outcomes explored in this study such as ability to practice compassion, job satisfaction, and burnout. Finally, we also note that, in relation to compassion, this study focuses on the clinicians’ outcomes. We call for future research to triangulate these findings with patients-related outcomes and patients and families’ experiences of compassion.

### Conclusions

The most important contribution of this study lies in providing robust empirical evidence attesting to the effect working in value discrepant environments on compassion and other associated outcomes in a large sample of healthcare professionals. The ability to show compassion as well as outcomes associated with clinician wellbeing were all negatively and significantly impacted by discrepancies between individual and organisational values. Many, if not most, clinicians enter their profession with the desire to help others and feel confident about their compassion competency. However, they also progressively find that priorities of their organisations are inconsistent and in some incidences in conflict with their own values, hindering their ability to provide compassion. Put simply, working in healthcare is not how they envisioned, negatively affecting clinicians’ wellbeing and performance. In response, healthcare organisations, policy makers and government agencies must critically assess how operational processes, practices, performance indicators, and resource allocation models contribute to value discrepancies and begin to make changes that bring values into line with one another. Consonance between values will likely lead to better employee performance, professional wellbeing, and patient care and, consequently, lower healthcare costs and more efficient healthcare systems.

## Data Availability

Due to risk of possibility of clinicians reidentification, the data is not publicly available, but can be made available upon request under certain conditions

## Acknowledgments

We thank Greg West (Kai Tahu, Kati Mamoe), and Arianna Sutton (Kai Tahu, Kati Mamoe, Waitaha oku Iwi) who have provided their clinical expertise in designing this study; Greg West and Ariana Sutton were also our cultural advisors who helped to align this study to reflect Māori worldview and values. In addition, we would like to thank FMHS Responsiveness to Māori team at Waipapa Taumata Rau (The University of Auckland) – Dr Kimiora Henare (Te Rarawa, Te Aupōuri) and Dr Karen Wright (Ngāi Tahu) - who provided guidance with regards to the upholding to the Te Tiriti O Waitangi (The Treaty of Waitangi). We additionally thank Kiralee Schache and Amelie Tuato’o who have helped with the study recruitment, and Harrison Boss who provided feedback and guidance with regards to Sinclair Compassion Questionnaire (SCQ) factor analyses. Use of the Sinclair Compassion Questionnaire (SCQ) authored by Dr. Shane Sinclair, Dr. Tom Hack, Dr. Cara MacInnis, Harrison Boss, Priya Jaggi, Susan McClement, Aynharan Sinnarajah and Genevieve Thompson was made under license from UTI Limited Partnership. Finally, we thank all clinicians who took part in this study as well as organisational managers who have helped to distribute this study.

## Footnotes

• Contributors: AP has conceived and designed this study, conducted recruitment, analysis and data interpretation and drafted the manuscript. NC and SJP has supervised the study procedures and are the guarantors. All authors contributed to study design and the recruitment of participants. All authors equally contribute to data interpretation, reviewed, and approved the final version on this manuscript. The corresponding author attests that all listed authors meet authorship criteria and that no others meeting the criteria have been omitted.

• Funding: This study is a part of a PhD funded by Margaret Burland Scholarship at the University of Auckland. The funder was not involved in the design and conduct of the study; collection, analysis, interpretation of data, writing of the report, or decision to submit the article for publication.

• Competing interests: All authors have completed the ICMJE uniform disclosure form at www.icmje.org/coi_disclosure.pdf and declare no support from any organisation for the submitted work; no financial relationships with any organisations that might have an interest in the submitted work in the previous three years; no other relationships or activities that could appear to have influenced the submitted work.

• Ethical approval: This study was approved by the Auckland Health Research Ethics Committee on the 21st of October 2021 (Approval Number AH23221) and received locality approvals from each of 20 national District Health Boards (DHBs) in Aotearoa/New Zealand. Electronic informed consent was obtained from all participants before completing the survey

• Data sharing: Due to risk of possibility of clinicians reidentification, the data is not publicly available, but can be made available upon request under certain conditions.

• Dissemination to participants and related patient and public communities: The findings of this study will be disseminated to all participating District Health Boards and Unions. In addition, our media relation departments will plan to further disseminate through press releases, as well as our institutional websites. The authors of the study will utilise Twitter to distribute via individual accounts.

• The manuscript’s guarantors (NC, SJP) affirm that the manuscript is an honest, accurate, and transparent account of the study being reported; that no important aspects of the study have been omitted; and that any discrepancies from the study as planned (and, if relevant, registered) have been explained.

• The Sinclair Compassion Questionnaire Healthcare Provider Ability Self Assessment (HCPASA) and the Sinclair Compassion Questionnaire Healthcare Provider Competence Self Assessment (HCPCSA) are available at www.compassionmeasure.com or by emailing.

