## Appendix for "Working in values-discrepant environments inhibits clinicians’ ability to provide compassion and reduces wellbeing: a cross-sectional study"

### Appendices

#### Appendix 1: EFA figures for SQC-HCPASA scale

##### *Correlation table, screeplot and parallel analysis*

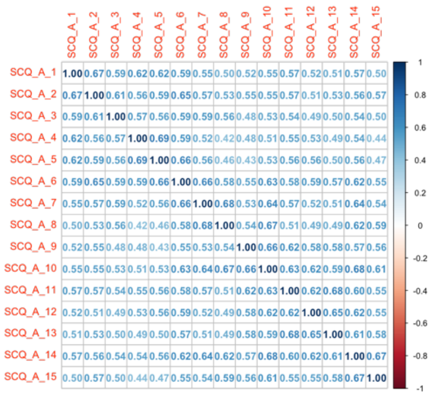


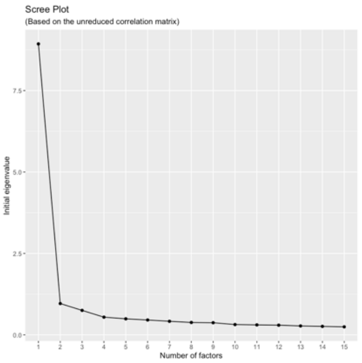


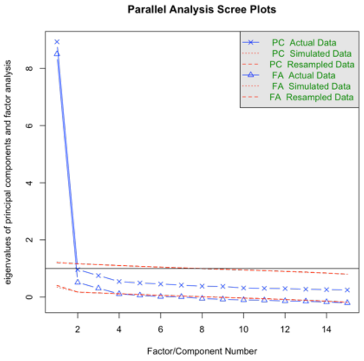


##### *Factor loadings*

| | ***Item wording*** | **Factor loadings** |
|---|---|---|
| SCQ_A_1 | *Feel cared for* | 0.75 |
| SCQ_A_2 | *Genuine concern* | 0.76 |
| SCQ_A_3 | *Communicating is a sensitive manner* | 0.72 |
| SCQ_A_4 | *Attentive* | 0.71 |
| SCQ_A_5 | *Providing comfort* | 0.74 |
| SCQ_A_6 | *Very supportive* | 0.80 |
| SCQ_A_7 | *Providing care* | 0.77 |
| SCQ_A_8 | *Speaking with kindness* | 0.72 |
| SCQ_A_9 | *Seeing as person* | 0.72 |
| SCQ_A_10 | *Behaving in caring way* | 0.80 |
| SCQ_A_11 | *Really understanding* | 0.78 |
| SCQ_A_12 | *Good relationship* | 0.74 |
| SCQ_A_13 | *Seeing patients' perspective* | 0.74 |
| SCQ_A_14 | *Warm presence* | 0.80 |
| SCQ_A_15 | *Sincere* | 0.73 |

Appendix 2: EFA for SQC-HCPCSA scale

*Correlation table, screeplot and parallel analysis*

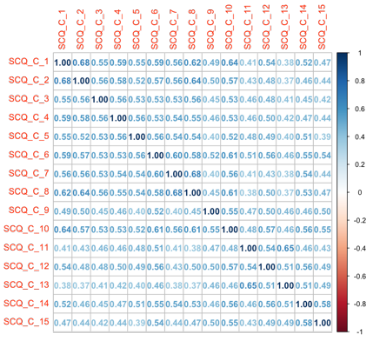

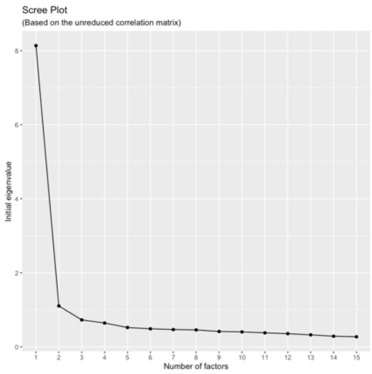


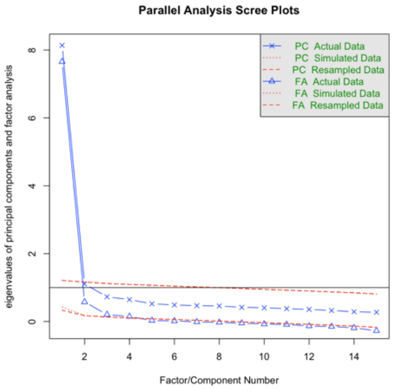


*Factor loadings*

| | ***Item wording*** | **Factor loadings** |
|---|---|---|
| SCQ_C_1 | *Feel cared for* | 0.77 |
| SCQ_C_2 | *Genuine concern* | 0.74 |
| SCQ_C_3 | *Communicating is a sensitive manner* | 0.70 |
| SCQ_C_4 | *Attentive* | 0.72 |
| SCQ_C_5 | *Providing comfort* | 0.70 |
| SCQ_C_6 | *Very supportive* | 0.78 |
| SCQ_C_7 | *Providing care* | 0.72 |
| SCQ_C_8 | *Speaking with kindness* | 0.76 |
| SCQ_C_9 | *Seeing as person* | 0.66 |
| SCQ_C_10 | *Behaving in caring way* | 0.78 |
| SCQ_C_11 | *Really understanding* | 0.65 |
| SCQ_C_12 | *Good relationship* | 0.71 |
| SCQ_C_13 | *Seeing patients' perspective* | 0.62 |
| SCQ_C_14 | *Warm presence* | 0.72 |
| SCQ_C_15 | *Sincere* | 0.66 |

### Appendix 3: PPOVD_HCS scale derivation of values

| | Values based on those signalled by healthcare institutions | Values that predict compassion or lack of thereof in healthcare (systematic review) | Values emphasized by senior Māori clinicians | Value emphasized by non-Māori clinicians | Values missing from the perspective of PVQ |
|---|---|---|---|---|---|
| Partnership: to work in partnership with people | ■ | ■ | ■ | ■ | |
| Excellence: to value excellence | ■ | ■ | | | |
| Compassion: to treat people with compassion | ■ | ■ | ■ | ■ | |
| Hospitality: to provide hospitality and comfort to people | | | ■ | | |
| Objectivity: to value objectivity | | ■ | | | |
| Respect: to show respect towards people whoever they are | ■ | | ■ | ■ | |
| Culture: to value people's culture and customs | | | ■ | | |
| Holistic care: to value holistic care | | | ■ | | |
| Equity: to value equity | ■ | ■ | | | |
| Relationships: to establish good relationships with people | ■ | | ■ | | |
| Humanity: to recognise the humanity inherent in each person | ■ | | | | |
| Control: to value control | | | ■ | | |
| Personal autonomy: to value personal autonomy | | ■ | | | |
| Creativity: to value creativity | | | | ■ | |
| Integrity: to value integrity | ■ | | | | |
| Stability: to value stability | | | | | ■ |
| Safety: to value safety | ■ | | | ■ | |
| Spirituality: to value spirituality | | | ■ | | |
| Efficiency: to value efficiency | | ■ | | | |
| Enjoyment: to value enjoyment | | | | | ■ |
| Authority: to recognise the value of authority | | | ■ | | |
| Professional image: to value professional image | | | | | ■ |
| Money: to value money | | ■ | | | |

#### Appendix 4: EFA figures for PPOVD_HCS scale

##### *Correlation table*

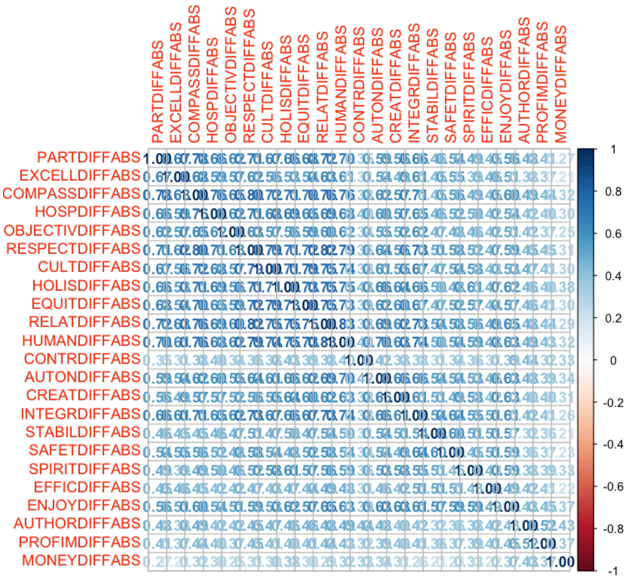


##### *Screeplot and parallel analysis*

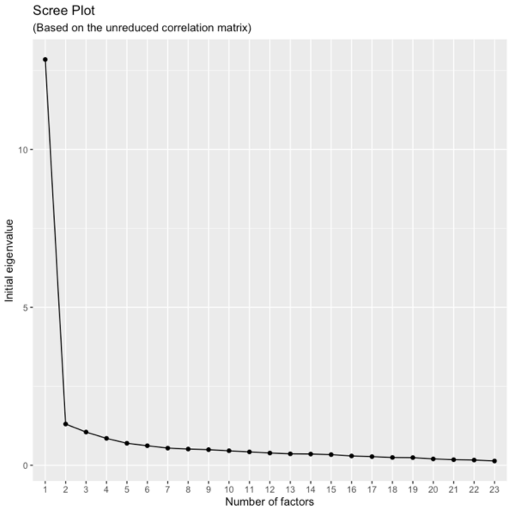


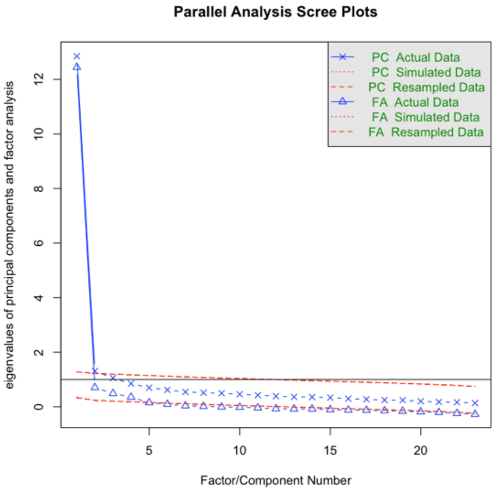

*Factor models*

| | 2 Factor model factor loadings | | 1 Factor model factor loadings |
|---|---|---|---|
| | Factor 1 | Factor 2 | Factor 1 |
| Respect | **0.96** | -0.09 | **0.86** |
| Compassion | **0.95** | -0.09 | **0.85** |
| Relationships | **0.86** | 0.04 | **0.88** |
| Culture | **0.84** | -0.01 | **0.82** |
| Partnership | **0.84** | -0.03 | **0.80** |
| Humanity | **0.81** | 0.09 | **0.87** |
| Hospitality | **0.79** | 0.02 | **0.79** |
| Equity | **0.77** | 0.07 | **0.82** |
| Excellence | **0.73** | 0.00 | **0.71** |
| Integrity | **0.72** | 0.13 | **0.83** |
| Objectivity | **0.65** | 0.09 | **0.72** |
| Holistic care | **0.64** | 0.22 | **0.83** |
| Authority | 0.01 | **0.64** | **0.58** |
| Money | -0.18 | **0.64** | 0.40 |
| Professional image | -0.02 | **0.63** | **0.54** |
| Enjoyment | 0.21 | **0.61** | **0.75** |
| Control | -0.01 | **0.56** | **0.49** |
| Spirituality | 0.23 | **0.50** | **0.67** |
| Efficiency | 0.16 | **0.49** | **0.59** |
| Stability | 0.22 | **0.47** | **0.64** |
| Creativity | 0.34 | **0.45** | **0.73** |
| Autonomy | 0.01 | **0.42** | **0.78** |
| Safety | 0.40 | 0.32 | **0.68** |

### Appendix 5: Correlation table

| | 1* | 2 | 3* | 4* | 5* | 6 | 7* | 8* | 9* | 10* | 11* | 12* | 13 | 14 | 15 | 16 | 17 | 18 | 19 | 20 | 21 | 22 | 23 | 24 | 25 |
|---|---|---|---|---|---|---|---|---|---|---|---|---|---|---|---|---|---|---|---|---|---|---|---|---|---|
| 1 Gender[a]* | 1 | | | | | | | | | | | | | | | | | | | | | | | | |
| 2 Age | -0.067* | 1 | | | | | | | | | | | | | | | | | | | | | | | |
| 3 Māori* | 0.037 | -0.016 | 1 | | | | | | | | | | | | | | | | | | | | | | |
| 4 Pākehā* | 0.049 | 0.119*** | -0.463*** | 1 | | | | | | | | | | | | | | | | | | | | | |
| 5 Tauiwi (Non-Māori/non-Pākehā)* | -0.081** | -0.117*** | -0.275*** | -0.725*** | 1 | | | | | | | | | | | | | | | | | | | | |
| 6 Years of experience | -0.057 | 0.832*** | -0.130*** | 0.149*** | -0.061* | 1 | | | | | | | | | | | | | | | | | | | |
| 7 Occupation[b]* | 0.215*** | -0.046 | 0.130*** | -0.009 | -0.092** | -0.177*** | 1 | | | | | | | | | | | | | | | | | | |
| 8 Urban vs. rural setting[c]* | -0.103*** | 0.078* | -0.013 | -0.043 | 0.057 | 0.092** | -0.063* | 1 | | | | | | | | | | | | | | | | | |
| 9 Orgnaisational sized[d]* | -0.049 | -0.059 | -0.055 | 0.048 | -0.009 | -0.026 | -0.053 | -0.159*** | 1 | | | | | | | | | | | | | | | | |
| 10 Private vs. public funding[e]* | 0.096** | -0.06 | -0.015 | -0.054 | 0.070* | -0.047 | 0.05 | -0.033 | -0.334*** | 1 | | | | | | | | | | | | | | | |
| 11 Care setting* | -0.081** | -0.090** | -0.078* | 0.002 | 0.058 | -0.072* | -0.115*** | -0.044 | 0.315*** | -0.147*** | 1 | | | | | | | | | | | | | | |
| 12 Belonging to cultural framework[g]* | 0.044 | 0.047 | 0.091** | -0.128*** | 0.068* | -0.005 | 0.090** | 0.023 | 0.090** | -0.121*** | 0.132*** | 1 | | | | | | | | | | | | | |
| 13 Patient numbers seen per week | -0.112*** | -0.03 | -0.033 | -0.036 | 0.066* | -0.005 | -0.171*** | 0.068* | -0.096** | 0.064* | 0.046 | 0 | 1 | | | | | | | | | | | | |
| 14 Hour or work per week | -0.250*** | -0.122*** | 0.052 | -0.103*** | 0.072* | -0.069* | -0.198*** | 0.049 | 0.122*** | -0.069* | 0.136*** | 0.075* | 0.041 | 1 | | | | | | | | | | | |
| 15 Absenteeism (in days) | 0.092** | -0.073* | 0.169*** | -0.067* | -0.058 | -0.139*** | 0.219*** | -0.077* | 0.014 | -0.01 | -0.006 | 0.113*** | -0.004 | 0.01 | 1 | | | | | | | | | | |
| 16 Intention to retire early (in years) | 0.016 | -0.253*** | -0.084** | -0.038 | 0.107*** | -0.159*** | -0.070* | -0.018 | 0.001 | 0.058 | 0.042 | -0.01 | 0.067* | -0.048 | -0.009 | 1 | | | | | | | | | |
| 17 Value discrepancy | -0.070* | 0.017 | 0.047 | 0.083** | -0.127*** | 0.054 | -0.072* | -0.029 | 0.132*** | -0.112*** | -0.013 | -0.201*** | -0.063* | 0.023 | 0.015 | 0.029 | 1 | | | | | | | | |
| 18 Job Satisfaction | 0.028 | 0.043 | 0.064* | -0.003 | -0.047 | 0.007 | 0.027 | 0.028 | -0.04 | 0.021 | -0.057 | 0.014 | -0.063* | -0.028 | -0.092** | -0.172*** | -0.359*** | 1 | | | | | | | |
| 19 Burnout | 0.059 | -0.236*** | -0.038 | 0.048 | -0.023 | -0.166*** | -0.071* | 0.013 | 0.106*** | -0.028 | 0.108*** | 0.012 | 0.104** | 0.098** | 0.146*** | 0.180*** | 0.229*** | -0.511*** | 1 | | | | | | |
| 20 Compassion competency | 0.214*** | 0.144*** | 0.099** | 0.025 | -0.104** | 0.111*** | 0.151*** | -0.107** | 0.036 | -0.018 | -0.046 | 0.074* | -0.068* | -0.135*** | 0.055 | -0.05 | 0.016 | 0.032 | -0.136*** | 1 | | | | | |
| 21 Compassion ability | 0.126*** | 0.178*** | 0.06 | -0.053 | 0.011 | 0.109** | 0.213*** | -0.072* | -0.081* | 0.057 | -0.129*** | 0.094** | -0.076* | -0.110*** | 0.024 | -0.070* | -0.266*** | 0.314*** | -0.387*** | 0.488*** | 1 | | | | |
| 22 Compassionate love | 0.153*** | 0.002 | 0.122*** | -0.065 | -0.024 | -0.04 | 0.136*** | -0.046 | -0.022 | 0.009 | -0.036 | 0.012 | -0.033 | -0.088** | 0.061 | -0.062 | -0.084* | 0.100** | -0.134*** | 0.314*** | 0.270*** | 1 | | | |
| 23 Fears of compassion towards others | -0.04 | -0.201*** | -0.082* | -0.067* | 0.136*** | -0.139*** | -0.025 | 0.067* | -0.053 | 0.080* | 0.034 | 0.113*** | 0.083* | 0.006 | 0.085* | 0.127*** | -0.098** | -0.161*** | 0.285*** | -0.205*** | -0.134*** | -0.198*** | 1 | | |
| 24 Self-efficacy | 0.06 | 0.181*** | 0.023 | 0.056 | -0.079* | 0.165*** | 0.099** | -0.039 | 0.043 | 0.062 | -0.022 | 0.05 | -0.01 | -0.051 | 0.007 | -0.095** | -0.06 | 0.225*** | -0.244*** | 0.371*** | 0.349*** | 0.154*** | -0.141*** | 1 | |
| 25 Social desirability | 0.085* | 0.123*** | 0.017 | -0.054 | 0.046 | 0.097** | 0.078* | -0.061 | -0.03 | -0.045 | 0.024 | 0.123*** | -0.019 | -0.038 | -0.03 | -0.029 | -0.134*** | 0.108** | -0.232*** | 0.252*** | 0.294*** | 0.150*** | -0.174*** | 0.262*** | 1 |

a Gender (0=male, 1=female). b occupation (1=doctor, 2 = nurse, 3 = allied health). c urban vs. rural (1=urban, 2=rural). d organisational size (0=small/medium, 1=large). e private vs. public (1=public, 2=private). f setting (1=primary, 2=secondary/tertiary). g belonging to cultural framework (0=no, 1=yes)

*Pearson's correlation coefficient, unless denoted (*) - Spearman Rank correlation coefficient*
